# Exercise Interventions for Rheumatoid Arthritis: A Sequential Bibliometric and Content Analysis — An Evidence Mapping Study

**DOI:** 10.64898/2026.05.27.26354187

**Authors:** Zihan Zou, Ziyi Zhang, Ran Zhao, Yanjun Liu, Jingqi Gao, Lei Gu

**Author notes:** Corresponding author.Hunan University of Chinese Medicine,No.300,Xueshi Road,Yuelu District,Changsha,China.Email address (Lei Gu). These authors contributed equally to this work (Zihan Zou, Ziyi Zhang).

## Abstract

**Background:** This two-stage sequential study combined bibliometric data with standardised full-text coding to build a modality-outcome evidence matrix for RA exercise research, testing whether publication hotspots align with directionally consistent clinical evidence. Conventional bibliometric studies highlight hotspots and collaboration patterns but cannot determine whether heavily discussed topics rest on clinically coherent trial evidence. The gap between thematic popularity and empirical consistency has been noted, but few studies have quantified it systematically.

**Methods:** We searched WoSCC and PubMed for RA exercise studies (2016–2025). Three nested datasets were defined: Dataset A (n=352) for bibliometric mapping; Dataset B (n=203) for full-text coding; and Dataset C (n=54) for evidence-matrix synthesis. Using CiteSpace and VOSviewer, we assessed publication trends, collaboration patterns and keyword bursts, and coded each trial for exercise type, outcome domain and effect direction. For directional consistency, we used an 80% threshold as a pragmatic cut-off, with sensitivity tests at 70% and 85% (Supplementary Table S6). RoB 2.0 and GRADE were not performed; the study was not PROSPERO-registered—both are explicit limitations.

**Results:** Publications rose from 24 (2016) to 49 (2025). Keyword bursts shifted from cardiovascular risk (2016–2019) to fatigue (2020–2022) to quality of life (2023–2025). Aerobic and resistance training showed the highest evidence concentration (≥15 studies/outcome) and directional consistency (≥85%). Mind-body exercise had moderate volume (n=16) but weaker consistency (60-70%), largely due to heterogeneous protocols. HIIT, BFRT, cardiovascular risk, and body composition remain underexplored (≤5 studies/outcome).

**Conclusion:** This descriptive evidence map shows aerobic and resistance training are the most thoroughly studied modalities; mind-body and novel interventions need standardised protocols and larger trials. Our framework distinguishes research visibility from evidence coherence. The matrix summarises volume and directional agreement only—not a clinical guideline or comparative-effectiveness assessment.

## 1. Introduction

Rheumatoid arthritis (RA) is a chronic autoimmune disease characterised by synovitis, progressive joint destruction, and systemic inflammation ^[1]^, affecting approximately 0.5%–1.0% of the population and impacting multiple organs beyond joints ^[2][3]^. Although disease - modifying antirheumatic drugs (DMARDs) and biologic agents are the mainstay of treatment, long-term use is associated with adverse effects that may lead to discontinuation, resulting in inadequate disease control and reduced quality of life ^[4]^. Thus, identifying safe and effective non - pharmacological adjunctive exercise interventions is clinically important. Exercise has gained increasing recognition as a core component of comprehensive RA management. However, whether research hotspots reflect accumulated evidence or merely publication volume remains unclear.

Previous studies have shown that regular exercise improves muscle strength, joint function, and cardiorespiratory fitness without exacerbating disease activity ^[5]^; however, the comparative effects of different modalities and their optimal parameters remain unclear ^[6]^. The 2018 EULAR recommendations for physical activity in people with inflammatory arthritis and osteoarthritis advocate for regular aerobic and strengthening exercise as part of standard care ^[7]^. As of the search date (February 2, 2026), these 2018 recommendations remain the most comprehensive physical activity guidance specifically for RA; we therefore use them as the primary comparator. Nonetheless, these guidelines recommend exercise types based on expert consensus and existing systematic reviews without quantifying the consistency of evidence across different exercise–outcome pairs. Several meta-analyses have reported that aerobic exercise improves cardiorespiratory fitness and reduces disease activity in RA ^[8]^, and that resistance training improves muscle strength and physical function ^[9]^. However, traditional meta - analyses primarily pool effect sizes and cannot distinguish whether a research topic is merely a publication hotspot or is supported by directionally consistent evidence. Evidence mapping, defined as a systematic approach to visualise the distribution, volume, and consistency of research in a given field, offers a middle ground between bibliometric description and meta - analytic synthesis ^[10]^. Unlike meta - analysis, which pools effect sizes and requires protocol homogeneity, evidence mapping catalogues research activity and directional patterns without assuming comparability for statistical pooling. Similar integrated bibliometric and content analysis frameworks have been applied in other chronic disease fields such as stroke rehabilitation and diabetes self - management, demonstrating the feasibility of combining publication trend analysis with clinical evidence consistency evaluation.

Despite the growing body of literature, an important gap remains. Most existing reviews have focused on individual exercise modalities. A recent bibliometric study by Xu et al. (Medicine, 2023) summarised publication trends, collaboration networks and research hotspots over the past two decades ^[11]^. However, Xu et al.’s approach had three specific limitations: (1) it relied exclusively on publication metadata without extracting full-text intervention protocols; (2) it did not classify exercise modalities or outcome domains systematically; and (3) it could not determine whether identified hotspots (e.g., fatigue, quality of life) were supported by directionally consistent clinical findings. For example, “fatigue” emerged as a strong keyword burst after 2020, yet no one has systematically checked whether different exercise modalities produce consistent effects on fatigue across trials. Existing single- stage bibliometric tools analyse publication metadata but do not include standardised full-text trial coding. Our sequential design therefore maps the global research landscape first and then quantifies cross-trial outcome consistency, precisely to resolve the mismatch between how often a topic is discussed and how coherent the underlying evidence actually is. We believe this distinction matters because volume does not equal maturity.

We hypothesised that bibliometric hotspots—identified by keyword bursts—would show variable directional consistency across modalities, and that some clinically important domains (cardiovascular risk, body composition) would remain evidence-sparse despite thematic visibility.

To fill this gap, we built an analytical framework that combines bibliometric mapping with structured content analysis in sequence. The bibliometric part identified thematic clusters and temporal trends; the content-analysis part then assessed internal coherence, outcome consistency and evidence concentration within those clusters ^[12]^. The resulting evidence - distribution matrix gives a visual summary of where the studies are concentrated and whether their findings point in the same direction for each exercise–outcome pair ^[13]^.

Our study had three main aims. First, we aimed to describe the temporal evolution, national/institutional collaboration patterns and keyword hotspots of RA exercise research from 2016 to 2025 using bibliometric tools. Second, and more centrally, we quantified the volume and directional consistency of evidence across all exercise–outcome pairs by coding 203 original trials and building an evidence matrix. Third, we tested whether bibliometric hotspots are a good proxy for coherent clinical evidence, and in doing so, offered a transferable analytical template for similar mapping studies in other fields.

## 2. Methods

### 2.1 Research Framework

The study followed a two-stage sequential design. In the first stage, bibliometric mapping was conducted to identify publication trends, collaboration patterns, and thematic structures. In the second stage, structured full-text content analysis was applied to original intervention studies to examine intervention characteristics, outcome distribution, and directional consistency. The coded data were then synthesised into an evidence matrix. This framework was guided by directed qualitative content analysis, in which coding categories were predefined and refined during pilot coding. Ethics approval was not required because the study was based exclusively on previously published literature retrieved from public databases and did not involve human participants, animal subjects, or primary data collection, in accordance with institutional guidelines for secondary research.

To avoid ambiguity, three nested datasets were predefined and used consistently throughout the manuscript:

Dataset A (bibliometric dataset): all eligible records included in bibliometric analysis only (n=352).

Dataset B (intervention coding dataset): original exercise intervention studies extracted from Dataset A for full-text coding (n=203).

Dataset C (evidence-matrix dataset): a subset of Dataset B that provided sufficiently standardised exercise protocols and extractable outcome reporting for modality-outcome synthesis (n=54).

Dataset A included all 352 eligible records for bibliometric mapping. Dataset B was derived from Dataset A by restricting to 203 original intervention studies (excluding reviews, meta-analyses, protocols, and non-interventional studies). Dataset C was a further subset of Dataset B (n=54) with sufficiently standardised exercise protocols for evidence-matrix synthesis. Dataset C included only single-modality interventions (aerobic, resistance, hand exercise, mind-body, HIIT, aquatic, walking, whole-body vibration); multimodal interventions (n=28 in Dataset B) were excluded from Dataset C because their inseparable exercise components prevented meaningful cross-study consistency assessment. Dataset definitions are summarised in Supplementary Table S1.

The overall research workflow, including the two-stage sequential design and the construction of the three nested datasets, is illustrated in Fig. 1.

**Fig. 1.** Research framework and literature analysis flow diagram. The two- stage sequential design is shown: Stage 1 (bibliometric mapping, Dataset A, n=352); Stage 2 (full- text coding, Dataset B, n=203); and evidence- matrix synthesis (Dataset C, n=54).

### 2.2 Search strategies

We searched the Web of Science Core Collection (WoSCC) and PubMed on February 2, 2026. All references cited in this manuscript were published before that date. WoSCC provides standardised citation data essential for co-authorship and co-citation analyses; PubMed was added to capture broader biomedical literature. The Cochrane Library was excluded because its primary content consists of systematic reviews and protocols without the standardised citation metadata required for bibliometric network analysis (co-authorship, co-citation, keyword co-occurrence). Embase was excluded because its citation metadata export format is not fully compatible with CiteSpace’s required input format, and including it would have required substantial manual reformatting. This decision may have reduced coverage of non-WoSCC-indexed trials, which we acknowledge as a limitation. Records published in January–February 2026 were excluded because their publication window was less than one month, which is insufficient for meaningful bibliometric indexing and citation analysis.

WoSCC search: TS=(exercise OR physical activity OR physical exercise OR sport OR sports training) AND TS=(rheumatoid arthritis OR RA).

PubMed search: (Exercise[Mesh] OR Motor Activity[Mesh] OR Sports[Mesh] OR exercise[Title/Abstract] OR exercises[Title/Abstract] OR physical activity[Title/Abstract] OR physical activities[Title/Abstract] OR physical exercise[Title/Abstract] OR sports[Title/Abstract] OR sports training[Title/Abstract]) AND (Arthritis, Rheumatoid[Mesh] OR rheumatoid arthritis[Title/Abstract] OR RA[Title/Abstract])

Initial retrieval yielded 6,909 records. Exclusions: 1,769 non-original/reviews; 2,935 outside 2016–2025; 25 non-English. After removing 33 duplicates identified using EndNote’s duplicate detection function (based on title, author, and journal matching) followed by manual verification, 2,147 records remained. Title and abstract screening then excluded 1,795 records (non-interventional studies: n=1,203; mixed arthritis populations with non-extractable RA data: n=327; non-exercise interventions: n=265), resulting in 352 records eligible for bibliometric analysis (Dataset A). Of these, we further identified 203 original intervention studies (RCTs, quasi-experimental, and pre-post controlled trials) for content analysis (Dataset B); the remaining 149 records (reviews, meta-analyses, protocols, and non-interventional studies) were retained for bibliometric mapping but excluded from the evidence matrix. From Dataset B, 54 studies (Dataset C) met the additional criteria for inclusion in the evidence matrix. Fig. 2 presents the complete PRISMA-style screening flow.

**Fig. 2.** PRISMA - style flowchart of literature screening (February 2, 2026). Dataset A: bibliometric analysis (n=352); Dataset B: content coding (n=203); Dataset C: evidence matrix (n=54). Reasons for exclusion at the title/abstract screening stage are shown: non-interventional studies (n=1,203); mixed arthritis populations without separable RA data (n=327); non-exercise interventions (n=265).

### 2.3 Inclusion and exclusion criteria

#### Bibliometric analysis (Dataset A)

All publication types (original articles, reviews, protocols, editorials) indexed in WoSCC/PubMed within 2016–2025, English-language, related to RA and exercise/physical activity.

#### Content analysis (Dataset B)

Original intervention studies (RCTs, quasi- experimental, pre- post) evaluating structured exercise or physical activity interventions in RA patients, reporting clinical, behavioural, or mechanistic outcomes. Exclusion: animal studies, conference abstracts, non- original publications, mixed populations without separable RA data, non- exercise interventions.

#### Evidence matrix (Dataset C)

Studies from Dataset B with sufficiently explicit and replicable exercise protocols (type, frequency, intensity, duration), outcomes assignable to predefined domains, and extractable directional findings. Multimodal interventions with inseparable components were excluded.

Two reviewers (Z. Zou and Z. Zhang) independently screened all records. Inter-rater reliability was assessed on a 50- article sample (κ=0.834), representing 24.6% of Dataset B. Disagreements occurred in 6.2% of screening decisions (n=22 out of 352) and were resolved by discussion, with a third reviewer (L. Gu) adjudicating unresolved cases.

### 2.4 Bibliometric Analysis

Bibliometric analyses were performed using CiteSpace ^[14]^ and VOSviewer ^[15]^, with a time span of 2016–2025 and 1-year slices. Node types included countries, institutions, authors, and keywords. For co-occurrence analysis, terms appearing at least five times were included using association strength normalisation. The g-index (k=25) was selected per CiteSpace official standard for medical bibliometric research to balance high-yield papers and minor publications (Chen, 2004) . Burst detection used default parameters (γ=1.0); network pruning used the Pathfinder algorithm to simplify redundant network nodes; clustering used the log-likelihood ratio (LLR) algorithm. Co-occurrence threshold ≥5 times follows common medical visualisation norms. Publication trends were visualised using simple bar charts rather than predictive polynomial fitting, because retrospective publication data should not be used for forecasting ^[16]^.

### 2.5 Data Extraction

Excel 2021 was used for basic statistical analyses. A standardised extraction form was designed, piloted on 15 randomly selected articles, and refined before full extraction. Extractor training included two calibration sessions with the pilot articles; discrepancies were resolved through consensus coding before independent extraction commenced. The overall analysis covered annual publication volume, collaboration networks among countries, institutions, and authors, as well as keyword clustering and burst detection. The extracted variables included the first author, publication year, exercise type, intensity, frequency, session duration, and intervention period. Clinical outcomes were categorised as “improved,” “worsened,” or “unchanged” based on reported changes in measures such as the Disease Activity Score in 28 joints (DAS28), visual analogue scale (VAS), Health Assessment Questionnaire (HAQ), the 6- minute walk test (6MWT), fatigue, and quality of life. Data on physiological mechanisms were also extracted, including inflammatory factors, signalling pathways, and immune cells. For studies with incomplete reporting in these areas, the corresponding variables were coded as “not addressed.” Further details are provided in the Supplementary Materials.

Directionality was coded hierarchically: when MCID thresholds were available, they were used as the primary criterion for classifying improvement/no change/worsening (DAS28 reduction ≥0.6 or achieving low disease activity; VAS reduction ≥10 mm on a 100- mm scale or ≥1 point on a 10- point scale; HAQ reduction ≥0.22; 6MWT increase ≥30 m). When MCID data were unavailable, author- reported statistical significance (p<0.05) was used as a secondary criterion. When both were available and conflicted, MCID- based classification took precedence.

### 2.6 Content analysis

Full-text structured coding was performed for all studies in Dataset B (n=203). Coding used a hybrid approach: categories were predefined and refined after pilot coding of 15 articles. Two independent coders randomly selected 50 intervention articles from the whole dataset for double-blind coding; Cohen’s κ was 0.834, representing almost perfect consistency (Supplementary Figure S1).. All coding disagreements were discussed; unresolved divergences (n=12, 5.9% of Dataset B) were resolved by group discussion among the two coders and the corresponding author, with majority rule (≥2 of 3) applied, according to a pre-defined unified coding manual (Supplementary File S1).

Exercise types were classified into nine primary categories (Fig. 9): aerobic (n=92), resistance (n=30), multimodal (n=28), hand exercise (n=20), mind-body (n=16), HIIT (n=8), aquatic (n=5), walking exercise (n=2), and whole-body vibration (n=1). An additional category of “non-exercise co-interventions” (n=149) comprised physical activity counselling, behavioural modification, and dietary interventions; these were not classified as exercise modalities and were excluded from the evidence matrix but retained for descriptive bibliometric analysis.

Classification rules for multimodal interventions: If a study explicitly combined modalities as a single arm (e.g., aerobic + resistance), it was coded as “multimodal.” If independent arms were reported separately, each was coded independently. If a study reported both aerobic and resistance components but did not explicitly designate a primary modality, we classified it as “multimodal” only if the authors described it as such or if both components were given equal emphasis in the Methods section; otherwise, we coded by the dominant component based on session frequency or duration. Ambiguous cases were resolved by group discussion.

Outcomes were grouped into seven domains: disease activity, pain, physical function, patient-reported outcomes, cardiovascular risk, body composition, and inflammatory markers. Multiple indicators within the same domain were not merged; consistent results were recorded as a single finding, conflicting results as inconsistent. Among the 203 studies, 49 were RCTs; their design features are summarised in Supplementary Table S2.

### 2.7 Evidence Matrix Classification Criteria

We used the following criteria to build the evidence matrix shown in Fig. 10. A threshold of five or more studies per outcome pair was chosen to define a “recurrently studied” category, following the approach used in earlier evidence-mapping work ^[10]^. For directional consistency, we set an 80% cut-off as a practical working rule. We also assessed the robustness of our classification by testing alternative minimum study thresholds (≥3, ≥5, ≥8 studies per outcome pair). The overall classification remained stable across these thresholds (>90% of modality – outcome pairs stayed in the same category; Supplementary Table S3). We want to be open with readers: this 80% value is not derived from GRADE or from any formal consensus standard. It is a pragmatic choice, and we chose it mainly to avoid over-interpreting results from very small or heterogeneous sets of trials. This threshold follows the precedent set by Miake-Lye et al. (2016) in their evidence-mapping methodology, where a 75-80% consistency cut - off was recommended for classifying “directionally consistent” findings. To see whether our conclusions depended heavily on this particular number, we re-ran the classification using 70% and 85% thresholds and summarised the results in Supplementary Table S5. For pairs with eight or more studies, the classification stayed stable (>90% of modality – outcome pairs remained in the same category). For pairs with only five to seven studies, changing the threshold from 70% to 85% could change the classification from “consistent” to “mixed” for 1-2 marginal pairs; these are explicitly noted in the discussion.

Before going further, we should clarify three descriptive terms that appear throughout the paper. By “evidence concentration” we simply mean the number of studies available for a given exercise – outcome pair. “Directional consistency” is the percentage of those studies that report effects going in the same direction—whether that direction is improvement, no change or worsening. When we use the phrase “evidence coherence” in the discussion, we mean a pair that has at least five studies and shows 80% or greater directional agreement. These are purely descriptive labels. They do not imply anything about study quality, effect size, statistical precision or GRADE-level certainty. We have tried to use these terms consistently so that readers can see exactly what we are measuring.

For inclusion in the evidence matrix (Dataset C), a study had to meet three conditions: (1) the exercise prescription was reported clearly enough to be replicable (type, frequency, intensity and duration); (2) the outcome could be assigned to one of our seven pre- defined domains; and (3) we could confidently extract the directional finding (improved / unchanged / worsened) from the full text. We excluded multimodal interventions where the exercise components could not be separated, and we also excluded non-exercise co-interventions such as counselling or dietary advice. These exclusions were made to keep the matrix reasonably homogeneous; without a minimum level of protocol standardisation, cross-study comparison of directional consistency would not be meaningful.

A crucial methodological limitation needs to be stated here. This study was not prospectively registered in PROSPERO or any similar public repository ^[17]^. That absence increases the risk of post-hoc analytic choices. The absence of pre-registration may have increased analytic flexibility in coding rule refinement and threshold selection, potentially inflating apparent consistency if coding decisions were influenced by observed data patterns. We discuss this openly in Section 4.6. We also did not conduct formal risk - of - bias assessment using Cochrane RoB 2.0, nor did we apply GRADE grading^[18–19]^. As a result, the matrix and Table 5 reflect only the volume and directional consistency of the available studies, not a judgement of their methodological quality or the certainty of the evidence. Any clinical interpretation drawn from this matrix should therefore be seen as a descriptive summary of what the literature looks like, not as a practice guideline.

### 2.8 Risk of Bias and Evidence Certainty (Declared Limitations)

We acknowledge that this study did not perform a formal risk- of- bias assessment using the Cochrane RoB 2.0 tool for the 49 RCTs, nor did we apply GRADE (Grading of Recommendations, Assessment, Development and Evaluations) to determine the certainty of evidence. Therefore, the evidence matrix and the summary in Table 5 are based solely on study volume and directional consistency, without weighting for methodological quality. Without quality weighting, the matrix may overrepresent evidence from methodologically weak studies. For example, if the majority of mind- body exercise studies are low quality but consistently positive, the matrix would classify them as “consistent” without quality adjustment. This limitation means that our findings should be interpreted as a descriptive overview of the research landscape rather than a definitive guide for clinical decision- making. Future updates of this evidence map should integrate RoB 2.0 assessments and GRADE grading to provide a more robust evidence synthesis.

## 3. Results

### 3.1 Bibliometric Results

#### 3.1.1 Publication Output and Temporal Trends

The annual publication trend is shown in Fig. 3. From 2016 to 2025, the number of publications on exercise-related research in RA showed an overall upward trend, despite several short-term fluctuations. Annual output increased from 24 in 2016 to 49 in 2025, with temporary declines observed in 2020 and 2024. The most pronounced growth occurred after 2020, reaching a peak of 54 publications in 2023. This upward trend may reflect the increasing recognition of exercise as an important component of RA management, together with a growing emphasis on non- pharmacological interventions, functional outcomes, and quality of life. Nonetheless, publication volume alone does not necessarily reflect evidence quality; this should be interpreted in conjunction with the evidence distribution shown in Fig. 10.

**Fig. 3.** Annual publication output trend from 2016 to 2025. The quadratic fitting model is used only for historical data description and cannot predict future publication volume. (x represents sequential years starting from 2016).

#### 3.1.2 National contributions and international collaboration

Country-level collaboration is presented in Table 1 and Fig. 4. The United States had the highest publication output (n=70) and a centrality of 0.77. Brazil, Sweden, the United Kingdom, and China followed with 38, 33, 33, and 27 publications, respectively. Canada showed a centrality of 0.42, followed by France (0.25), Sweden (0.19), and China and the Netherlands (both 0.17). By contrast, countries such as Turkey and Brazil showed relatively low centrality despite moderate publication output. The collaboration network was centered on the United States, with the United Kingdom, Sweden, China, Brazil, and the Netherlands forming the main connected nodes. Overall, international collaboration in this field was concentrated in a limited number of countries, mainly in North America and Western Europe.

**Fig. 4.** Country collaboration network (generated by VOSviewer, threshold ≥5 publications).

#### 3.1.3 Institutional Research Output

According to the top 10 institutions ranked by publication volume (Table 2), the most productive institutions included the University of Copenhagen, Leiden University Medical Center, Copenhagen Center for Arthritis Research, and several related clinical or academic units. Fig. 5 shows that the institutional collaboration map was dominated by a small number of major centers, with the Department of Rheumatology, Harvard Medical School, the University of Birmingham, the University of Copenhagen, and Karolinska University Hospital occupying prominent positions. The typical year for the leading institutions was mainly 2016, indicating that their core contributions appeared early in the study period. For some institutions, such as the University of Birmingham and related clinical units, the typical year was 2017, whereas it was 2018 for the University of Copenhagen- related nodes. These results suggest that institutional output was concentrated in a limited number of centers and that the periods of active publication varied across institutions.

**Fig. 5.** Institutional collaboration network (CiteSpace, g-index k=25).

#### 3.1.4 Author Research Output

According to the publication output of authors listed in Table 3, Hamilton, Roschel had the highest number of publications (n=12), followed by Kitas, George D. and Bruno, Gualano (n=11 each), and Metsios, George S. and Mannerkorpi, Kaisa (n=9 each). Duda, Joan L., Fenton, Sally A., and Appel, Bente Esbensen each contributed eight publications, while Alexander, Fraser had seven. In the author collaboration network based on edge weights (Fig. 6A), Kitas, George D., Metsios, George S., Hamilton, Roschel, Bruno, Gualano, and Duda, Joan L. occupied relatively central positions and showed strong links with other authors. The co-citation network (Fig. 6B) showed that Kitas, George D., Metsios, George S., Duda, Joan L., and Fenton, Sally A. were among the most frequently cited authors, with larger nodes and denser connections. The color distribution suggests that earlier active authors were mainly concentrated in the 2016–2019 period, whereas more recent contributions were more visible after 2020.

**Fig. 6.** Author maps (6A)Author collaboration network (VOSviewer, association strength normalization). (6B)Author co-citation network (VOSviewer, association strength normalization).

#### 3.1.5 Journals and Co- cited Journals

As shown in Table 4,Rheumatology Internationalhad the highest publication output (n=30 articles) among the top journals.Arthritis Care & Research,Arthritis Research & Therapy, and several other rheumatology-related journals also appeared frequently, indicating that this topic was mainly published in journals in rheumatology, rehabilitation, and clinical medicine. In the journal network map (Fig. 7A),Rheumatology International,Arthritis Care & Research, andArthritis & Rheumatologyoccupied relatively central positions and showed dense connections with other journals. The dual-map overlay (Fig. 7B) illustrates the intellectual flow of the field. The citing literature (right hemisphere) is predominantly sourced from clinical medicine journals (e.g., rheumatology, rehabilitation medicine), represented by the orange path labeled “Medicine, Medical, Clinical.” The cited literature (left hemisphere) draws heavily from molecular biology, immunology, and basic science journals (green path labeled “Molecular, Biology, Immunology”). Additionally, the yellow path (“Health, Nursing, Medicine”) and the blue path (“Psychology, Education, Health”) indicate secondary connections to nursing, public health, and behavioral sciences. This overall pattern suggests that research on exercise interventions in RA is built upon a strong mechanistic foundation, bridging basic immunology with clinical rehabilitation and behavioral health.

**Fig. 7.** Journal maps (7A)Journal co - occurrence network (CiteSpace).k (7B)Dual - map overlay of journals (CiteSpace).

#### 3.1.6 Keywords analysis of research hotspots

Keyword timeline and clustering analyses (Figs. 8A and 8C) revealed “physical activity” (#0) as the largest and most sustained cluster. Earlier clusters (“physical exercise,” “disease activity,” “inflammatory arthritis”) were associated with disease status and exercise intervention, whereas later clusters (“quality of life,” “mobile applications,” “hand function,” “chronic pain,” “patient reported outcome measures,” “patient involvement”) indicated a shift toward patient- centered and rehabilitation-related topics.

**Fig. 8.** Keyword maps. (8A) Keyword timeline view. (8B) Keyword density map. (8C) Keyword clustering map. (8D) Strongest citation bursts (Dataset A, CiteSpace [6.4.R1], g- index k=25).Strength = citation burst intensity.

**Fig. 9.** Distribution of exercise intervention types among the 352 included records.

**Fig. 10.** Evidence matrix bubble chart of exercise modality vs. clinical outcome. Bubble size reflects study count, color reflects directional consistency. Neither dimension reflects study quality or effect size. Color indicates directional consistency: green, ≥80% of studies reporting improvement; yellow, mixed (neither direction ≥80%); red, ≥80% no change or worsening. Gray bubbles denote <5 studies (insufficient evidence).

The density map (Fig. 8B) showed “rheumatoid arthritis,” “physical activity,” “disease activity,” and “physical exercise” as the highest- density core, surrounded by “quality of life,” “fatigue,” “pain,” and “rehabilitation.”

Burst detection (Fig. 8D) revealed a progression from early bursts (“patient perspective,” “inflammatory arthritis,” “oxidative stress”) to later bursts (“fatigue,” “cardiorespiratory fitness,” “aerobic exercise,” “systematic review”). However, citation bursts reflect rising attention rather than evidentiary consistency, and thus do not necessarily indicate conclusive findings.(Strength = citation burst intensity; full data shown in Supplementary Table S4.)

#### 3.1.7 Cross-Stage Comparison: Bibliometric Hotspots Versus Evidence-Matrix Consistency

We next checked whether the topics that stood out in the keyword bursts also showed coherent evidence in the matrix. This is not a formal statistical comparison, because the two stages operate at different levels—keyword bursts capture topic-level attention in the wider literature, while the matrix reflects intervention - outcome directional agreement among the subset of trials that had codable protocols (Dataset C). Still, looking across the two layers, we noticed three instructive mismatches.

Fatigue had the strongest keyword burst after 2020 (strength = 4.56, 2020 – 2022). But when we looked at specific modality-outcome pairs, the consistency varied a great deal: aerobic exercise studies reported improvements in fatigue in 85% or more of cases (12 studies), whereas mind - body studies showed only 63% agreement (8 studies). So a strong bibliometric signal at the keyword level did not translate into a uniform picture at the intervention level. Cardiovascular risk, meanwhile, showed an early burst (2016 – 2019) but turned up in fewer than five studies for most exercise modalities — thematic prominence without much empirical density. Body composition followed a similar pattern: the keyword has attracted growing attention, yet the actual number of studies for each modality remains very small (12 studies total across all modalities, with only 7 reporting improvement, 58.3%). Taken together, these cases suggest that a bibliometric hotspot is not a reliable proxy for evidence coherence. This is precisely the kind of dissociation that a single-stage bibliometric study would miss. We caution readers, however, that this comparison is descriptive pattern recognition, not a formal interaction test.

### 3.2 Exercise Prescription Dimension: Distribution and Characteristics of Intervention Modalities

The bibliometric analysis above identified major thematic clusters and keyword bursts. To examine whether the evidence within these clusters is actually consistent and mature, we next performed content analysis on the 203 original intervention studies (Dataset B). Fig 9 shows the distribution of intervention categories among the 352 records, separating exercise modalities from non-exercise co - interventions (e.g., counselling, dietary advice). Importantly, as shown in the evidence matrix (Fig. 10), a high volume of studies does not always correspond to high consistency. For example, while “fatigue” emerged as a keyword burst after 2020, the evidence matrix reveals that its directional consistency varies markedly across exercise modalities.

#### 3.2.1 Overall Distribution of Exercise Intervention Modalities

Among the 203 original intervention studies, 54 (26.6%) delivered structured exercise interventions with clearly defined protocols. Fig. 9 shows the distribution of exercise modalities among these studies. Aerobic training (n=92, 45.3%) and resistance training (n=30, 14.8%) together accounted for 60.1% of all original intervention studies in Dataset B. Other categories included multimodal (n=28, 13.8%), hand exercise (n=20, 9.9%), mind- body (n=16, 7.9%), HIIT (n=8, 3.9%), aquatic (n=5, 2.5%), walking exercise (n=2, 1.0%), and whole- body vibration (n=1, 0.5%). The “non- exercise co-interventions” category (n=149, 73.4%) comprised physical activity counselling, behavioural modification, and dietary interventions without standardised exercise protocols; these were excluded from the evidence matrix but retained in bibliometric analysis.

#### 3.2.2 Traditional Exercise Modalities: Aerobic and Resistance Training

Aerobic exercise currently has the strongest evidence base among the various exercise modalities. Most studies have shown that aerobic exercise improves VO₂peak, six-minute walking distance, and DAS28 without exacerbating joint symptoms ^[8]^. However, evidence regarding the safety of high-intensity aerobic exercise in patients with active RA remains insufficient. Resistance training has been reported to be superior to aerobic training in improving rectus femoris thickness, lean body mass, and functional test performance ^[9]^. Nevertheless, long-term follow-up data for both of these traditional intervention modalities remain limited.

#### 3.2.3 Mind-Body and Alternative Exercise

Yoga has been shown to reduce disease activity, Health Assessment Questionnaire Disability Index (HAQ-DI) scores, and several inflammatory markers, with depressive symptoms decreasing in a time-dependent manner ^[20]^. Follow-up findings further suggest that participants in yoga interventions are more likely to be employed full-time ^[21]^. One study reported that tai chi combined with hand exercises improved DAS28, erythrocyte sedimentation rate (ESR), C-reactive protein (CRP), and scores for depression and anxiety ^[22]^. However, some meta-analyses have indicated that tai chi does not produce statistically significant improvements in pain or physical function ^[23]^. Compared with conventional exercise modalities, mind-body exercises may offer the advantages of low impact and high acceptability; however, their optimal intensity and frequency remain unclear.

#### 3.2.4 Novel Physical Adjunctive Interventions

A few other modalities have drawn research interest, but their evidence base remains small. High-intensity interval training (HIIT) has been tested in a handful of studies; one 12-week supervised programme combined with strength training improved maximal oxygen uptake, grip strength and sit-to-stand performance ^[24]^, with some benefits sustained at 12 months ^[25]^. Blood-flow-restriction training (BFRT) and whole-body vibration have also been explored in preliminary work ^[26–29]^. However, in our Dataset C, all of these modalities had five or fewer studies for each outcome pair. Their safety in patients with active disease (DAS28 > 5.1) has not been adequately examined, and most of the existing trials are small. We have therefore moved the detailed discussion of these novel approaches to Section 4.4.1 (Future Research Directions), where they can be considered as priorities for feasibility and dose-finding work rather than as established options.

#### 3.2.5 Associations Between Exercise Modalities and Clinical Outcomes

The exercise modality–outcome patterns shown in Fig. 10 indicate that the available evidence is unevenly distributed across outcome domains. In this matrix, bubble size represents the number of studies for each exercise–outcome pair, and color represents directional consistency (green: ≥80% studies reporting improvement; yellow: mixed, with neither direction reaching ≥80%; red: ≥80% no change/worsening; gray: <5 studies). Aerobic exercise and resistance training account for most of the literature on core symptoms, physical function, and patient-reported outcomes, whereas mind-body exercise contributes a relatively substantial proportion of studies focused on patient-reported outcomes. In contrast, water-based exercise and hand exercises are less frequently represented. Blood flow restriction training, HIIT, and whole-body vibration training remain at the periphery of the evidence base, with relatively few studies addressing cardiovascular risk, inflammatory markers, or body composition. Overall, the current literature suggests that evidence for cardiovascular risk and body composition outcomes remains limited, highlighting the need for future studies with longer follow-up and broader outcome assessment. The number of studies within each modality–outcome pair reflects research concentration rather than comparative efficacy; a larger number of studies does not necessarily indicate superior clinical benefit.

Well - supported: Aerobic and resistance training showed ≥ 15 studies/outcome and ≥ 85% consistency across physical function, fatigue, and quality of life (Fig. 10, large green bubbles). Inconsistent: Mind-body exercise (16 studies) showed only 60-70% consistency across psychological outcomes (moderate - sized yellow bubbles). Evidence - sparse: Cardiovascular risk and body composition had ≤5 studies across all modalities (gray bubbles); HIIT, BFRT, whole-body vibration had ≤8 studies total.

#### 3.2.6 Design characteristics of included RCTs

Supplementary Table S6. presents the key design features of the 49 RCTs stratified by exercise modality. Behavioural/psychological interventions had the largest number of studies (n=11) and the largest median sample size (n=122 participants), but also the lowest outcome consistency (72.7%). Aerobic and resistance training showed high consistency (≥85.7%) with moderate sample sizes. Hand/local exercises achieved 100% consistency, albeit with short intervention durations (median 4 weeks). Aquatic exercise, whole-body vibration, and other novel modalities were represented by only one or two RCTs, limiting generalisability. Across modalities, RCTs with larger sample sizes (≥50 participants) tended to show lower consistency rates than smaller trials, a pattern that may reflect publication bias toward positive findings in small studies.

### 3.3 Evolution of Research Priorities

Keyword timeline and burst analyses identified three major phases in research development:

2016–2019: Safety, Disease Activity, and Cardiovascular Risk. During this period, exercise research in RA focused primarily on whether physical activity was safe in the context of inflammatory joint disease. Disease activity measures such as DAS28 and cardiovascular risk markers featured prominently. This pattern likely reflected longstanding concerns that exercise might exacerbate inflammation and the simultaneous recognition of cardiovascular disease as a major cause of mortality in RA.

2020–2022: Fatigue and Patient-Reported Outcomes. As advances in pharmacologic therapy improved control of overt joint inflammation, research attention shifted toward persistent symptoms not fully addressed by medication, especially fatigue and broader patient-reported outcomes. Investigators increasingly examined quality of life, sleep, self-reported function, and mental health.

Since 2023: Precision, Stratification, and Long-Term Management. Recent literature has emphasised individualised intervention design, risk stratification, and long-term quality of life. This reflects a broader transition from symptom suppression toward integrated long-term disease management, with growing interest in identifying which modality works best for which patient and outcome domain.

### 3.4 Clinical Outcome Dimension: Evidence Distribution and Consistency

#### 3.4.1 Core Pathological Symptoms

##### Disease Activity

DAS28 was the most frequently reported core pathological outcome. Most studies indicated that exercise does not increase disease activity, and several reported significant reductions ^[5][30]^. This finding is clinically important because it helps dispel the traditional fear that exercise may aggravate RA inflammation.

##### Pain

Pain reduction, commonly measured using the visual analogue scale (VAS), showed relatively consistent improvement across studies, with an overall moderate effect pattern. Although not all modalities were equally represented, the general trend supports exercise as a useful adjunctive strategy for pain management.

##### Morning Stiffness

Among the available modalities, combined aerobic plus resistance training showed the most pronounced effect on morning stiffness ^[31]^. Water-based exercise was also associated with shorter stiffness duration in some studies, though the evidence base was smaller and less mature.

#### 3.4.2 Physical Function and Body Composition

##### Functional Capacity

The six- minute walk test (6MWT) emerged as an important indicator of functional capacity. Aerobic exercise, resistance training, and combined training all significantly improved 6MWT performance, generally with moderate effect sizes. This supports the role of structured exercise in preserving mobility and endurance in RA.

##### Muscle Strength

Resistance training was frequently associated with improvement in muscle strength. This aligns with both physiological expectations and clinical needs, given the prevalence of weakness, deconditioning, and muscle loss in RA.

##### Grip Strength and Hand Function

Both hand- specific exercise programs and whole- body resistance training improved grip strength. Hand/local exercises, despite shorter intervention periods, showed highly consistent benefits, suggesting they may be especially useful when hand dysfunction is a prominent concern.

##### Body Composition

Evidence regarding body composition was less consistent. Aerobic exercise combined with dietary intervention reduced body fat percentage and waist circumference ^[32]^. However, the effect of exercise alone on body fat remained inconsistent across studies, reflecting both limited long- term data and variability in intervention intensity and measurement methods.

#### 3.4.3 Patient-Reported Outcomes

##### Fatigue

Fatigue was one of the most prominent patient-centred outcomes. Meta- analytic evidence suggests that aerobic exercise significantly reduces fatigue, though the degree of improvement may not always exceed the minimum clinically important difference (MCID) ^[33]^. A 12- week program combining HIIT with strength training improved multidimensional fatigue, and this effect persisted for up to 12 months in follow- up ^[34]^. Neuroimaging findings further suggest that fatigue improvement may be linked to structural connectivity involving the left insular cingulate cortex and the paracentral lobule ^[35]^.

##### Sleep

Walking- based interventions improved sleep quality, including Pittsburgh Sleep Quality Index scores, in some studies ^[36]^. However, the overall evidence remains limited and somewhat inconsistent ^[37]^.

##### Depression and Anxiety

Mind- body interventions such as yoga and tai chi, as well as mobile health- based self-management approaches, showed beneficial effects on depression and anxiety ^[38–40]^. These findings suggest that exercise in RA is not only biomechanical but also psychophysiological in effect.

##### Quality of Life

A meta- analysis reported an overall improvement in quality of life with a standardised mean difference of approximately 0.50, indicating a moderate beneficial effect ^[41]^. Quality of life gains appeared across several modalities, though the clearest support came from aerobic and mixed interventions.

#### 3.4.4 Risk of Systemic Complications

##### Cardiovascular Risk

Cardiovascular disease remains a leading cause of mortality in RA. A four-week aerobic exercise program significantly reduced cardiovascular risk and DAS28 while improving endothelial function ^[42]^. Other studies suggest that exercise may improve vascular function through mechanisms distinct from anti-TNF therapy ^[43]^. Exercise appears to show a possible dose-response trend in cardiovascular risk improvement.

Vascular Function and Cardiac Coupling: Supervised physical activity has been associated with improved vascular health and significantly enhanced ventricular-arterial coupling after two years ^[44]^. These findings indicate that exercise may contribute to long-term cardiovascular remodelling beyond symptom relief.

Sedentary Behaviour: Sedentary time was positively associated with 10-year cardiovascular disease risk, whereas light-intensity physical activity was negatively associated with this risk ^[45]^. This supports a broader clinical message: reducing sedentary behaviour may be beneficial even when moderate-to-vigorous exercise targets are difficult to achieve.

### 3.5 Physiological Mechanisms

This paper does not generate original animal or human molecular experimental data; the following is a synopsis of existing hypotheses from the literature.

Exercise is thought to exert anti-inflammatory effects through multiple pathways ^[46–49]^; myokines such as IL- 6, BDNF, and irisin may mediate muscle- brain crosstalk ^[50]^; improved autonomic function contributes to cardiovascular protection ^[51–53]^. Neuroimaging studies have linked fatigue improvement to structural connectivity changes in brain regions including the left insular cingulate cortex and the paracentral lobule ^[54]^. Acute aerobic exercise has also been shown to modulate serum BDNF levels in RA patients, with changes correlated with anxiety and depression scores ^[55]^. These remain hypothesis- generating in the RA context.

## 4. Discussion

### 4.1 Evolution of Research Priorities

Keyword evolution showed a clear trajectory: safety and cardiovascular risk (2016 – 2019) → fatigue (2020 – 2022) → quality of life and digital health (2023 – 2025). This progression parallels clinical developments: the 2016 EULAR cardiovascular risk recommendations ^[56]^ drove early cardiovascular focus; the 2023 EULAR fatigue guidelines ^[57]^ amplified patient - reported outcome research; and recent advances in digital rehabilitation technologies have enabled mobile health and remote monitoring studies.

However, the evidence matrix reveals that thematic evolution has not been uniformly matched by evidence accumulation. Fatigue became a major hotspot, yet exercise effects on fatigue varied substantially by modality—85% consistency for aerobic interventions but only 60-70% for mind-body exercise. This dissociation matters: fatigue is clinically important , but clinicians cannot assume that all exercise modalities are equally effective. Aerobic training has the strongest fatigue evidence; mind - body exercise remains promising but inconsistent. Similarly, cardiovascular risk— an early hotspot—remains evidence - sparse in the matrix, with ≤ 5 studies for most modalities despite RA’s elevated cardiovascular mortality. This gap between thematic visibility and evidentiary maturity has implications for research prioritisation and clinical decision-making.

### 4.2 Evidence Coherence Across Exercise Modalities

The evidence matrix shows a fairly polarised picture. Aerobic and resistance training account for the largest volume of evidence and show the highest directional consistency—85% or more of studies reporting improvement across core outcomes. Some of that volume may simply reflect a research preference for standardised, easily quantifiable interventions ^[7]^, but the consistency of findings across different settings and populations suggests a consistent signal that warrants further investigation in quality- weighted analyses (e.g., RoB 2.0- adjusted meta- analysis), rather than a pure publication artefact. However, because our matrix does not weight studies by quality, this interpretation should be treated as hypothesis- generating.

Multimodal combined interventions had a moderate number of studies, but their protocols varied so much in training duration, cycle length and component mix that we could not make meaningful cross-study comparisons. Mind- body exercise also had a moderate volume, but only 60- 70% of studies reported improvement in the same direction—probably because yoga and tai chi protocols differ widely in session frequency (weekly vs. daily), instructor qualifications (certified yoga therapists vs. general instructors), and cultural adaptation. The novel modalities (HIIT, aquatic exercise, whole- body vibration and neuromuscular activation) all remain at an early stage, with five or fewer studies per outcome pair. We discuss these specifically in Section 4.4.1, rather than giving them extended treatment here, precisely to avoid overstating their current evidentiary weight.

The large “non- exercise co-interventions” category (n=149) mainly reflects non- exercise co- interventions such as counselling and dietary advice. This finding reinforces a point made by earlier reviews ^[58]^: exercise- intervention research still lacks standardised reporting, and that limits what can be synthesised. In addition, most trials had short follow- up periods and concentrated on joint function and disease activity, leaving long-term data on body composition and cardiovascular outcomes surprisingly thin.

### 4.3 Methodological Contribution of the Two-Stage Framework

Compared with conventional CiteSpace- or VOSviewer-based bibliometric reports, the present two-stage framework offers three distinct methodological advantages.

First, it separates research visibility from clinical coherence. Bibliometric mapping alone can identify fast-growing topics, but full-text coding is needed to determine whether studies within those topics are methodologically comparable and directionally aligned. This distinction is particularly valuable in rehabilitation, where heterogeneous interventions often generate large literatures that are difficult to synthesise quantitatively. Xu et al. ^[11]^ identified hotspots using metadata only; our framework adds: (1) systematic classification of exercise modalities and outcome domains; (2) quantification of directional consistency across modality–outcome pairs; and (3) explicit cross-stage comparison of bibliometric hotspots versus evidence-matrix findings.

Second, it creates a pragmatic middle layer between bibliometrics and meta-analysis. When sufficient protocol homogeneity exists (e.g., similar exercise type, frequency, intensity, and outcome measures), meta-analysis is appropriate. When heterogeneity is substantial—as in mind-body exercise where yoga, tai chi, and qigong vary widely—evidence mapping provides a prior step to assess whether formal synthesis is justified. The evidence matrix reveals whether heterogeneity exists and whether formal synthesis is justified. In this study, the matrix showed that mind-body exercise is too heterogeneous for meta-analysis without standardisation, whereas aerobic and resistance training have sufficient consistency for comparative effectiveness research.

Third, it identifies false maturity signals—areas that appear active but are methodologically unstable. Fatigue is a prime example: a major hotspot, but with variable consistency across modalities.

### 4.4 Comparison with Prior Systematic Reviews and Guidelines

The 2018 EULAR physical- activity recommendations for inflammatory arthritis and osteoarthritis [7] encourage regular aerobic and strengthening exercise as part of standard care, and they emphasise individualised prescription. Our findings are broadly in line with that: aerobic and resistance training had the largest and most consistent evidence in our matrix. What our matrix adds is a quantitative picture of how much evidence underpins each modality–outcome pair. It shows, for example, that the support for aerobic and resistance training rests on a much larger and more consistent literature than the support for other modalities. Notably, the 2018 EULAR document did not specifically address mind- body exercise, HIIT or BFRT, which matches our observation that these areas are still poorly supported. We should also note that EULAR has updated its guidance in other domains—for instance, fatigue management in 2023 ^[57]^—and future updates of this evidence map should use the most current comprehensive recommendations when they become available.

Earlier meta- analyses have shown that aerobic exercise improves cardiorespiratory fitness and reduces disease activity, and that resistance training improves muscle strength and physical function. Our work is consistent with those conclusions, but it adds a layer of synthesis by showing that these effects are not only statistically significant but also directionally consistent across a large number of trials. At the same time, we found some divergence from previous reviews. For example, although some meta- analyses have suggested that mind- body exercise may help psychological outcomes ^[20][23]^, our matrix shows that the evidence is quite inconsistent across studies. That inconsistency is probably due to heterogeneity in intervention protocols (e.g. yoga style, session frequency, instructor qualifications) rather than to a lack of genuine effect. In our view, this means that standardising protocols should come before any further meta- analytic pooling.

#### 4.4.1 Future Research Directions for Modalities with Limited Evidence

The evidence matrix highlights several modalities that currently have five or fewer studies per outcome pair. These are not necessarily ineffective; they are simply under- studied, and they need a particular kind of attention in the next phase of research.

HIIT and BFRT should not yet be recommended for routine use. Existing studies have generally excluded patients with high disease activity (DAS28 > 5.1), so their safety profile in that subgroup is unknown. A sensible priority would be feasibility and safety studies first (e.g., a structured dose-escalation protocol with safety monitoring (e.g., DAS28 assessment every 4 weeks), followed by dose-finding work to establish intensity, frequency and session duration. Whole- body vibration has been examined in only one or two small studies ^[28][59],^ and the evidence is too preliminary even to judge efficacy. Researchers interested in this modality should start with pilot randomised trials that include a sham comparator. Aquatic exercise has a slightly larger literature, with buoyancy and thermal effects that may reduce joint loading ^[60]^ and improve pain and function, but the available trials are small and the outcome data are sparse. Larger, adequately powered RCTs are needed. Hand exercises are a distinct category because hand involvement is so common in RA. Programmes such as the SARAH protocol, nerve- mobilisation exercises and task- oriented training have shown promising results for grip strength, pinch strength and manual dexterity, and they can be delivered remotely via video or mobile applications. The consistency across the few existing trials is high (100% in the RCTs we coded), but the total number of participants is small, so confirmatory trials with larger samples should be a priority.

### 4.5 Clinical and Research Implications

For clinicians, Table 5 summarises the current literature in terms of evidence volume and directional consistency. We have deliberately removed evaluative language such as “better supported” or “promising” from this table, because without formal quality weighting or GRADE grading, those terms would be misleading. The table now describes what the literature contains, not what it recommends..For researchers, we see several high-priority gaps. Aerobic and resistance training no longer need more efficacy trials; they need protocol optimisation and long-term adherence studies. Mind-body interventions need standardised parameters before further trials are worthwhile. HIIT and BFRT need feasibility and safety work rather than large efficacy trials at this stage. And across all modalities, there is a shortage of tissue-level mechanistic studies—muscle biopsy or synovial-fluid work, for instance—that go beyond serum biomarkers. The EULAR guidelines recommend aerobic and strengthening exercise but do not specify optimal intensity, frequency or duration; those parameters remain poorly standardised across studies. Our matrix suggests that protocol heterogeneity is one of the main barriers to synthesis, especially for multimodal and mind-body interventions.

For health policy, the uneven evidence distribution implies that funding and guideline development should concentrate on well-studied modalities while also supporting early-phase exploration of novel ones. Journals and reviewers should recognise that a bibliometric “hotspot” is not the same as evidence readiness.

### 4.6 Strengths and Limitations

This study has several strengths. It combines bibliometric mapping with structured content analysis, so it can look at both research visibility and evidence coherence in the same project. The evidence matrix gives an intuitive visual summary of where studies are concentrated and how consistent their findings are across outcomes. And the framework is transferable: it could be used in other fields that deal with complex interventions and multidimensional outcomes, particularly those with heterogeneous protocols where meta- analysis is not feasible due to protocol heterogeneity.

We also need to be straightforward about the limitations. We group them into four categories.

#### First, search and selection bias

We limited the search to WoSCC and PubMed and included only English-language publications. The English- only restriction may exclude an estimated 5- 10% of relevant studies from non- English-speaking regions, potentially biasing the evidence matrix toward findings reported in English-language journals, which may overrepresent positive results (publication bias). That may have excluded relevant studies from regional journals or non- English sources. Future mapping studies should consider Embase or regional databases to improve coverage, and conduct language sensitivity analyses comparing English- only versus multilingual retrieval.

#### Second, coding and classification subjectivity

Even with dual independent coding and a Cohen’s κ of 0.834, classifying complex multimodal interventions and grouping outcomes into seven domains required judgment. Most disagreements (78%) were resolved by clarifying the classification rules for multimodal interventions. We tried to minimise this with a detailed coding manual, but some subjectivity remains. Future studies should develop more granular coding manuals with explicit decision trees for ambiguous cases.

#### Third, the absence of formal quality and certainty appraisal

We did not use Cochrane RoB 2.0 or GRADE. This is a major limitation. Without quality weighting, the matrix may overrepresent evidence from methodologically weak studies. For example, if the majority of mind- body exercise studies are low quality but consistently positive, the matrix would classify them as “consistent” without quality adjustment. The matrix shows volume and direction, not trustworthiness. We also did not register the protocol prospectively in PROSPERO, which leaves room for post- hoc analytical choices. Both points mean that our findings are descriptive, not evaluative, and should not be used as clinical guidance. Future updates of this evidence map should integrate RoB 2.0 assessments and GRADE grading.

**Fourth, the thresholds we chose**—five studies and 80% consistency—were pragmatic rather than data- driven. We ran sensitivity analyses at 70% and 85%, and for well-studied modalities the picture did not change much; but for pairs with five to seven studies, the classification could flip. Readers should keep that in mind. Evidence mapping also cannot compare effect sizes directly; the matrix shows concentration and consistency, not pooled efficacy. Network meta- analyses would be needed to establish comparative effectiveness. We also did not systematically assess health economics, implementation feasibility, patient adherence or long-term sustainability, so our findings have limited value for real- world implementation planning. Future work should incorporate those dimensions alongside evidence mapping.

## 5. Conclusions

Over the past decade, RA exercise research has moved from symptom control towards patient-centred outcomes—quality of life, fatigue and cardiovascular risk management. Aerobic and resistance training have the most concentrated and consistent evidence, while mind-body and novel modalities remain methodologically immature. A key message from our work is that publication volume is not a reliable proxy for evidence quality. The mismatch between bibliometric hotspots (fatigue, cardiovascular risk, body composition) and actual evidence consistency should caution researchers, reviewers and guideline developers against reading too much into citation bursts. However, these findings are descriptive and should be interpreted within the context of the study’s methodological limitations (absence of RoB 2.0 and GRADE, English-only retrieval, and pragmatic threshold choices).

The sequential combination of bibliometric mapping and structured content analysis offers a replicable template for other clinical areas with diverse interventions and uneven evidence, such as rehabilitation, chronic-disease self-management and mind-body medicine. This workflow is most suitable for fields with heterogeneous interventions and multidimensional outcomes where meta-analysis is not feasible due to protocol heterogeneity. It is not intended to replace formal systematic reviews or meta-analyses. The workflow—bibliometric clustering to identify thematic boundaries, followed by content coding and evidence-matrix construction—can help other researchers distinguish thematic visibility from evidentiary coherence, identify genuine gaps and direct resources towards areas with the greatest potential for clinical impact. Going forward, the field needs protocol standardisation, longer follow-up studies and formal quality appraisal to strengthen the evidence base for exercise interventions in RA.

## Supporting information

Supplementary materials including additional figures and tables

**Supplementary Materials:Supplementary materials.docx**

**Supplementary Table S1: Dataset architecture and analytical roles — includes definition of Dataset A/B/C, sample sizes, sources, and primary analytical purposes.**

**Supplementary Table S2: Key design characteristics of included RCTs stratified by exercise modality.**

**Supplementary Table S3: Sensitivity analysis of classification thresholds (≥3, ≥5, ≥8 studies).**

**Supplementary Table S4: Citation burst terms with strength values (top 15).**

**Supplementary Table S5: Clinical outcome evidence summary (full).**

**Supplementary Table S6: Study characteristics of the 49 randomized controlled trials included in the content analysis, grouped by exercise modality.**

**Supplementary Figure S1: Cohen’s kappa calculation output for inter-rater reliability.**

**Supplementary File S1: Complete coding manual for exercise modality and outcome domain classification.**

## Author Contributions

**Zihan Zou and Ziyi Zhang contributed equally to this work.**

**Zihan Zou: Conceptualisation, methodology, formal analysis, investigation, writing — original draft, visualisation.**

**Ziyi Zhang: Methodology, software, data curation, writing — review and editing, visualisation, validation of data extraction.**

**Ran Zhao: Software, formal analysis, data curation, writing—review and editing.**

**Yanjun Liu: Formal analysis, investigation, writing—review and editing.**

**Jingqi Gao: Data extraction, literature screening, evidence matrix construction, writing—review and editing, validation.**

**Lei Gu: Conceptualisation, supervision, project administration, writing — review and editing, funding acquisition.**

**All authors have read and agreed to the published version of the manuscript. Funding:University-level Research Project of Hunan University of Chinese Medicine:**

**Development Dilemmas and Advancement Strategies for the Integration of Physical Activity and Healthcare in Community- based Elderly Care under the New Era**

## Conflicts of Interest

The authors declare no conflicts of interest.

## Clinical trial registration number

Not applicable. This study is a bibliometric and content analysis of published literature and does not report the results of a clinical trial.

## Consent to Participate declarations

Not applicable. This study is a secondary analysis of published literature and does not involve direct human subject research.

## AI usage statement

Generative AI tools (specifically ChatGPT) were used only for language polishing and improving readability during the preparation of this manuscript. No AI tool was used to generate data, perform the bibliometric or content analyses, determine the scientific conclusions, or make interpretive decisions. All scientific content was reviewed and approved by the authors, who take full responsibility for the final version.

## Abbreviations

6MWT: 6- Minute Walk Test
DMARDs: Disease- Modifying Antirheumatic Drugs
ESR: Erythrocyte Sedimentation Rate
GRADE: Grading of Recommendations, Assessment, Development and Evaluations
HIIT: High- Intensity Interval Training
MCID: Minimal Clinically Important Difference
PROSPERO: International Prospective Register of Systematic Reviews
RA: Rheumatoid Arthritis
RoB: Risk of Bias
VAS: Visual Analogue Scale
WoSCC: Web of Science Core Collection

## Data Availability

Data availability statement: This study is a bibliometric and content analysis based on published literature retrieved from the Web of Science Core Collection. The search strategy, inclusion criteria, and extracted data are fully described in the manuscript and supplementary materials. No new primary data or code were generated. The raw bibliometric records can be accessed via Web of Science using the reported search query, and the coding framework is provided in the supplementary file.

## Notes

### Competing Interest Statement

The authors have declared no competing interest.

### Summary of Updates

Background: This two-stage sequential study combined bibliometric data with standardised full-text coding to build a modality-outcome evidence matrix for RA exercise research, testing whether publication hotspots align with directionally consistent clinical evidence. Conventional bibliometric studies highlight hotspots and collaboration patterns but cannot determine whether heavily discussed topics rest on clinically coherent trial evidence. The gap between thematic popularity and empirical consistency has been noted, but few studies have quantified it systematically. Methods: We searched WoSCC and PubMed for RA exercise studies (2016-2025). Three nested datasets were defined: Dataset A (n=352) for bibliometric mapping; Dataset B (n=203) for full-text coding; and Dataset C (n=54) for evidence-matrix synthesis. Using CiteSpace and VOSviewer, we assessed publication trends, collaboration patterns and keyword bursts, and coded each trial for exercise type, outcome domain and effect direction. For directional consistency, we used an 80 percent threshold as a pragmatic cut-off, with sensitivity tests at 70 percent and 85 percent (Supplementary Table S6). RoB 2.0 and GRADE were not performed; the study was not PROSPERO-registered-both are explicit limitations. Results: Publications rose from 24 (2016) to 49 (2025). Keyword bursts shifted from cardiovascular risk (2016-2019) to fatigue (2020-2022) to quality of life (2023-2025). Aerobic and resistance training showed the highest evidence concentration (15 or more studies per outcome) and directional consistency (85 percent or more). Mind-body exercise had moderate volume (16 studies) but weaker consistency (60 to 70 percent), largely due to heterogeneous protocols. HIIT, BFRT, cardiovascular risk, and body composition remain underexplored (5 or fewer studies per outcome). Conclusion: This descriptive evidence map shows aerobic and resistance training are the most thoroughly studied modalities; mind-body and novel interventions need standardised protocols and larger trials. Our framework distinguishes research visibility from evidence coherence. The matrix summarises volume and directional agreement only-not a clinical guideline or comparative-effectiveness assessment.

### Funding

- Hunan University of Chinese Medicine Institutional Research Project: A Study on the Development Challenges and Promotion Strategies of "Integration of Physical Fitness and Healthcare" in Community Elderly Care under the New Era Context.

