## Supplementary materials including additional figures and tables for "Exercise Interventions for Rheumatoid Arthritis: A Sequential Bibliometric and Content Analysis — An Evidence Mapping Study": Supplementary materials.docx

### **Supplementary File S1. Complete coding manual for exercise modality and outcome domain classification**

#### **S1.1 Purpose of the coding framework**

This appendix describes the coding rules used to classify exercise interventions and clinical outcomes in the evidence‑mapping component of the study. The aim of the framework was to improve consistency and transparency in assigning studies to intervention categories and outcome domains, while preserving the interpretability of heterogeneous RA exercise research.

#### **S1.2 General coding principles**

Coding was performed at the study level, with extraction based on the full text.

A study could contribute to multiple outcome domains, but only once to each modality–domain cell.

Exercise modality was classified according to the dominant active intervention component.

Studies involving multicomponent management were included in the modality‑specific map only when the exercise component was sufficiently explicit and could be meaningfully categorised.

When a study included more than one exercise arm, each arm was reviewed separately; however, for matrix visualisation, the study contributed to the modality category most directly aligned with the reported intervention structure.

If protocol reporting was too vague to assign a clear modality, the study remained in the broader intervention dataset (Dataset B) but was excluded from the standardised protocol evidence map (Dataset C).

#### **S1.3 Exercise modality definitions**

##### **S1.3.1 Aerobic exercise**

Studies were coded as aerobic exercise when the intervention primarily involved continuous or interval‑based activities intended to improve cardiorespiratory fitness, such as walking, treadmill exercise, cycling, dancing‑based aerobic training, or structured moderate‑to‑vigorous endurance exercise. Typical defining features included rhythmic whole‑body movement, repeated sessions over time, and explicit emphasis on endurance, fitness, or aerobic capacity.

##### **S1.3.2 Resistance training**

Studies were coded as resistance training when the intervention primarily aimed to improve muscular strength, endurance, or force production through external resistance or body‑weight loading, such as machine‑based strength training, free weights, elastic resistance bands, progressive resistance exercise, or strengthening programmes. If an intervention combined aerobic and resistance components but one mode clearly dominated the design and analysis, it was coded according to the dominant component; otherwise it could be considered multimodal.

##### **S1.3.3 Hand exercise**

Studies were coded as hand exercise when the intervention specifically targeted hand and/or wrist function, strength, mobility, dexterity, or symptom relief. This included hand strengthening, grip exercise, finger range‑of‑motion practice, task‑oriented hand rehabilitation, and home‑based hand exercise programmes.

##### **S1.3.4 Mind‑body exercise**

Studies were coded as mind‑body exercise when the intervention integrated physical movement with breath control, mental focus, relaxation, meditative practice, or postural awareness. Examples included yoga, Tai Chi, Qigong, and meditative movement programmes.

##### **S1.3.5 Aquatic exercise**

Studies were coded as aquatic exercise when the intervention was performed primarily in water, including aquatic aerobics, pool‑based strengthening, and hydrotherapy‑like exercise programmes emphasising structured movement.

##### **S1.3.6 High‑intensity interval training (HIIT)**

Studies were coded as HIIT when the intervention involved repeated short bouts of relatively high‑intensity exercise interspersed with recovery periods, and when the protocol explicitly aligned with interval‑based high‑intensity training principles.

##### **S1.3.7 Multimodal/combined exercise**

Studies were coded as multimodal exercise when the intervention intentionally combined two or more distinct exercise modes without a single dominant component, for example, aerobic plus resistance, flexibility plus strengthening plus balance, or comprehensive supervised rehabilitation exercise packages. **Important:**Multimodal interventions were included in Dataset B for descriptive purposes but excluded from Dataset C (evidence matrix) unless the exercise components could be confidently separated and each component independently assessed.

##### **S1.3.8 Walking‑based exercise**

Studies were coded as walking‑based exercise when walking was the central and explicitly prescribed intervention mode, rather than a minor element within a broader programme.

##### **S1.3.9 Whole‑body vibration**

Studies using vibration platforms as the principal physical intervention were coded as whole‑body vibration.

##### **S1.3.10 Neuromuscular or targeted activation exercise**

This category included targeted physical training focused on balance, coordination, motor control, joint stabilisation, or region‑specific activation when such programmes did not fit more conventional aerobic or resistance categories.

##### **S1.3.11 Non‑exercise co‑interventions**

This category (n=149) included physical activity counselling, behavioural modification, and dietary interventions without clearly defined, replicable exercise protocols. These were retained in Dataset A for bibliometric analysis but excluded from Dataset C. They were not classified as "exercise modalities" in the evidence matrix.

#### **S1.4 Outcome domain definitions**

##### **S1.4.1 Disease activity**

This domain included DAS28, CRP, ESR, swollen and tender joint counts where used as disease activity indicators, and composite inflammatory or immune‑related markers.

##### **S1.4.2 Pain**

This domain included pain measured using visual analogue scale (VAS), numeric rating scale (NRS), or other validated pain instruments.

##### **S1.4.3 Physical function**

This domain included objective or semi‑objective indicators of function and performance, such as HAQ‑related functional status, walking tests (6MWT), timed performance tasks, grip‑related function, joint mobility relevant to daily activity, and physical performance capacity.

##### **S1.4.4 Patient‑reported outcomes**

This domain included self‑reported constructs beyond core symptoms, such as health‑related quality of life (e.g., SF‑36, EQ‑5D), self‑efficacy, patient global assessment, psychological well‑being (depression, anxiety), perceived health status, fatigue, sleep quality, and participation‑related outcomes.

##### **S1.4.5 Cardiovascular risk / cardiometabolic outcomes**

This domain included outcomes related to cardiovascular fitness or metabolic risk, such as VO₂max or exercise capacity, blood pressure, lipid profile, arterial stiffness, endothelial indicators, and cardiometabolic risk markers.

##### **S1.4.6 Body composition**

This domain included body weight, BMI, fat mass, lean mass, muscle mass, waist circumference, and sarcopenia‑related or adiposity‑related indices.

##### **S1.4.7 Inflammatory markers**

This domain included inflammatory cytokines (TNF‑α, IL‑6, IL‑1β, etc.) and other systemic inflammatory biomarkers.

#### **S1.5 Rules for directional coding**

Each modality–outcome pairing was coded directionally as **improved**, **unchanged/mixed**, or **worsened**.

##### **S1.5.1 Improved**

A domain was coded as improved when the study reported a favourable post‑intervention effect for that domain, based preferably on between‑group comparison. Examples included reduction in pain or fatigue, improvement in functional test performance, reduction in DAS28, increase in aerobic fitness, or favourable change in body composition. MCID thresholds were used as the primary criterion when available.

##### **S1.5.2 Unchanged/mixed**

A domain was coded as unchanged/mixed when: no statistically significant effect was reported; findings within the same domain were inconsistent; some measures improved while others did not; or effect direction varied by time point or subgroup without a dominant pattern.

##### **S1.5.3 Worsened**

A domain was coded as worsened when the intervention was associated with an unfavourable change in the outcome domain. This category was expected to be uncommon.

#### **S1.6 Decision hierarchy for conflicting data**

When the result pattern was not straightforward, the following hierarchy was used:

Between‑group comparison over within‑group change

Primary endpoint over secondary endpoint

Post‑intervention endpoint over exploratory long‑term follow‑up

Composite clinical interpretation over isolated single‑measure fluctuation

Conservative classification as unchanged/mixed when ambiguity remained

**S1.7 Handling of multicomponent interventions**

If the study included education, counselling, digital support, behavioural coaching, or rehabilitation components in addition to exercise, the study was included in the evidence map only when the exercise prescription was sufficiently explicit and the exercise effect could be reasonably isolated. Otherwise, it remained in Dataset B for descriptive coding but not in Dataset C.

#### **S1.8 Examples of coding decisions**

**Example 1:** A supervised cycling programme performed three times per week for 12 weeks, reporting reduced DAS28 and improved VO₂peak.

**Modality:** Aerobic exercise

**Outcome domains:**Disease activity = improved; Cardiovascular risk = improved

**Example 2:**A hand rehabilitation programme improving grip strength but not changing quality‑of‑life score.

**Modality:**Hand exercise

**Physical function:**improved

**Patient‑reported outcomes:**unchanged/mixed

**Example 3:**A yoga programme with improved pain but mixed fatigue and no significant CRP change.

**Modality:**Mind‑body exercise

**Pain:**improved

**Fatigue:**mixed

**Inflammatory markers:**unchanged/mixed

| Document ID | Code A | Code B |
| --- | --- | --- |
| WOS:001649182900001 | 1 | 1 |
| WOS:001641595800001 | 1 | 1 |
| WOS:001628056500001 | 1 | 1 |
| WOS:001630853900001 | 1 | 1 |
| WOS:001623988900001 | 1 | 1 |
| WOS:001628004600047 | 2 | 4 |
| WOS:001609977600001 | 2 | 2 |
| WOS:001606213100001 | 4 | 4 |
| WOS:001607647000001 | 4 | 4 |
| WOS:001596124500001 | 1 | 1 |
| WOS:001591253800006 | 4 | 4 |
| WOS:001586091500010 | 4 | 4 |
| WOS:001575263300001 | 1 | 1 |
| WOS:001567171100003 | 3 | 3 |
| WOS:001568520600001 | 2 | 3 |
| WOS:001516587000001 | 3 | 3 |
| WOS:001482103600001 | 4 | 4 |
| WOS:001472799000001 | 4 | 4 |
| WOS:001451845200003 | 4 | 4 |
| WOS:001486953800001 | 4 | 4 |
| WOS:001437255400001 | 4 | 4 |
| WOS:001428155500001 | 1 | 1 |
| WOS:001427136000001 | 4 | 4 |
| WOS:001421692100001 | 2 | 1 |
| WOS:001411092000001 | 4 | 4 |
| WOS:001412946700001 | 4 | 4 |
| WOS:001408726900001 | 4 | 4 |
| WOS:001401065700001 | 4 | 4 |
| WOS:001566532000001 | 4 | 4 |
| WOS:001632870800018 | 1 | 1 |
| WOS:001389508100001 | 4 | 4 |
| WOS:001435416600025 | 4 | 4 |
| WOS:001585794400001 | 4 | 4 |
| WOS:001386969100001 | 4 | 4 |
| WOS:001379059100004 | 4 | 4 |
| WOS:001362812400001 | 3 | 1 |
| WOS:001368059300001 | 4 | 4 |
| WOS:001368190200001 | 4 | 4 |
| WOS:001342926600001 | 4 | 4 |
| WOS:001324062100001 | 2 | 2 |
| WOS:001312060800001 | 4 | 4 |
| WOS:001322955400001 | 4 | 4 |
| WOS:001298334100001 | 1 | 1 |
| WOS:001284592000001 | 3 | 3 |
| WOS:001261198200001 | 2 | 2 |
| WOS:001256966900001 | 4 | 4 |
| WOS:001234360800001 | 1 | 2 |
| WOS:001228169400001 | 4 | 4 |
| WOS:001501605800001 | 2 | 2 |
| WOS:001235699500008 | 4 | 4 |

2.Supplementary Figure S1

Supplementary Figure S1. Cohen’s kappa calculation output for inter‑rater reliability. Calculated using Lensym Cohen’s Kappa Calculator based on 50 randomly selected articles. Kappa = 0.834, observed agreement = 90.0%, expected agreement = 39.8%.

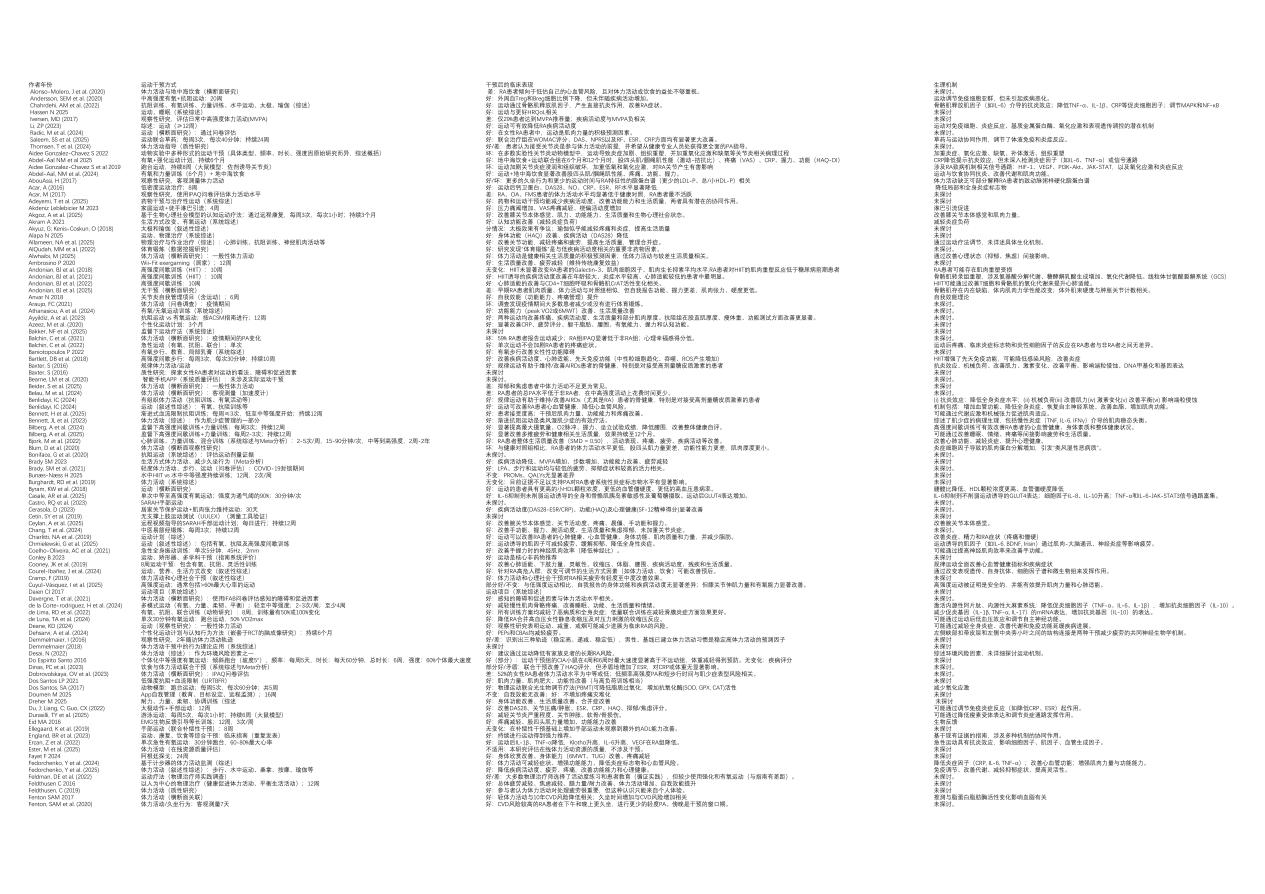

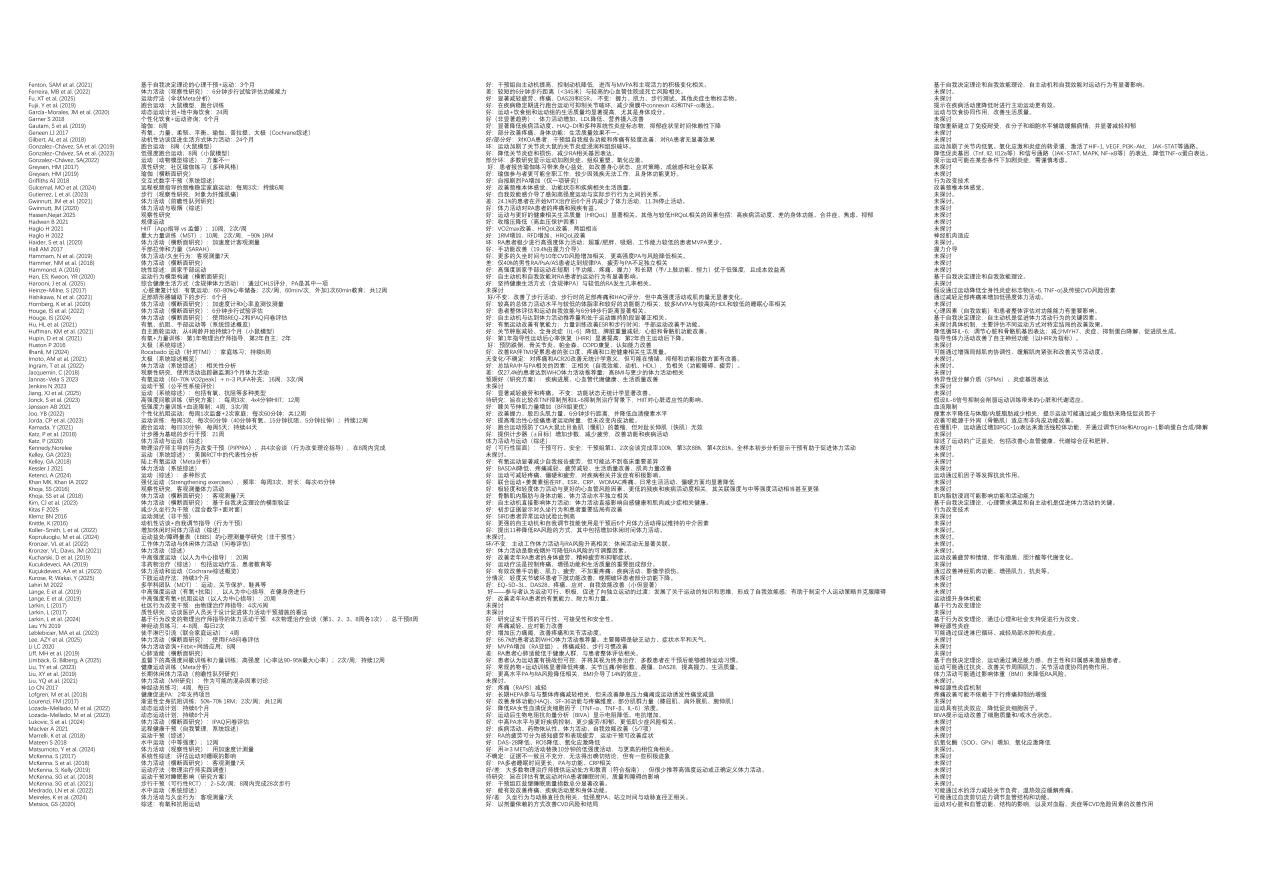

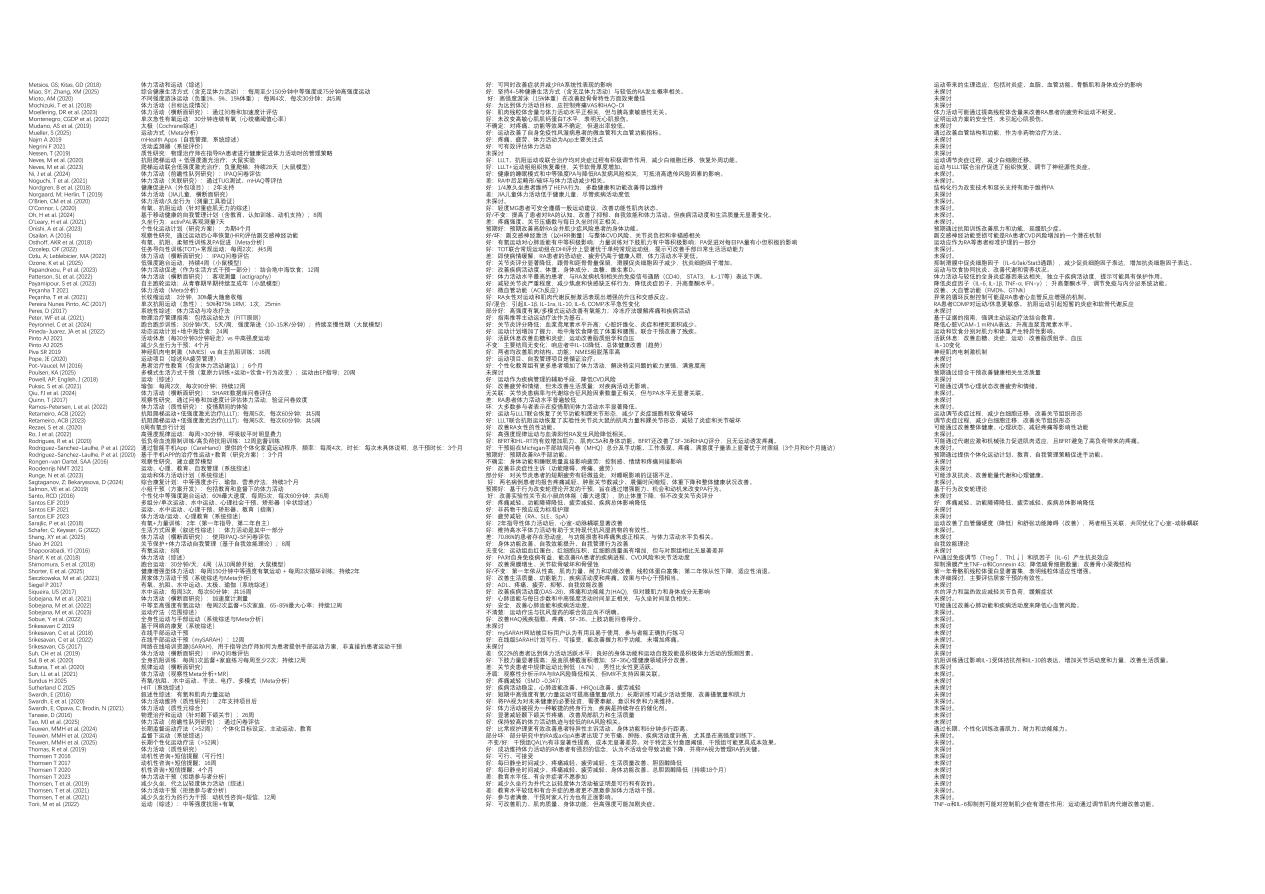

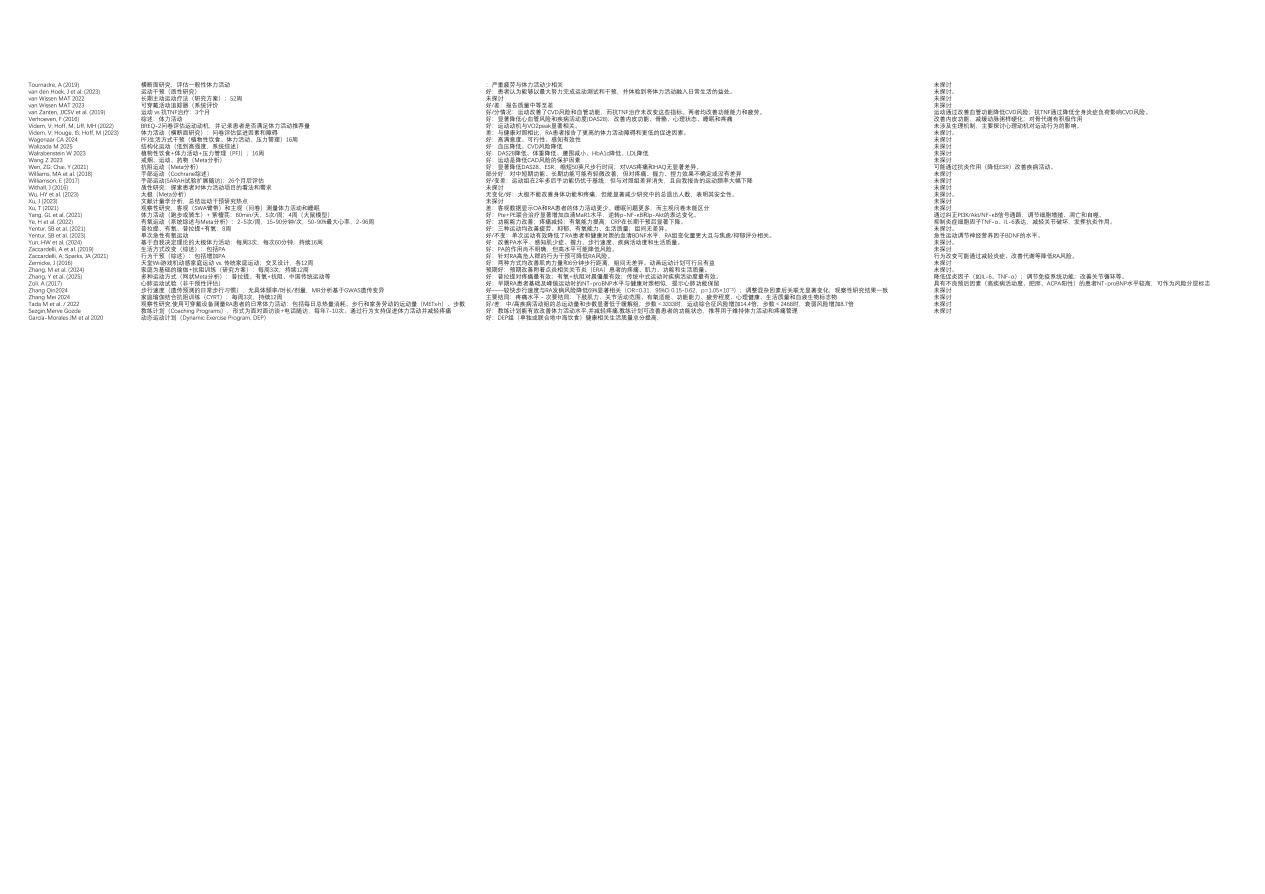
