## Supplementary materials including additional figures and tables for "Exercise Interventions for Rheumatoid Arthritis: A Sequential Bibliometric and Content Analysis — An Evidence Mapping Study": Supplementary Table S1.docx

Supplementary Table S1 Architecture of the included datasets and their analytical roles

| Dataset Level | Number of Records/Studies | Source and Scope | Main Inclusion Characteristics | Primary Analytical Purpose | Output in Current Study |
| --- | --- | --- | --- | --- | --- |
| Dataset A (Bibliometric dataset) | 352 | Records retrieved from WoSCC and PubMed, after deduplication and eligibility screening | English‑language publications related to rheumatoid arthritis and exercise/physical activity between 2016 and 2025; included original studies, reviews, meta‑analyses, protocols, editorials, and related scholarly outputs | To characterize annual publication growth, geographic collaboration, thematic evolution, keyword clusters, and citation bursts | Publication trend analysis; country/institution/author collaboration networks; keyword timeline, co‑occurrence, clustering, and burst analyses |
| Dataset B (Intervention coding dataset) | 203 | Full‑text screened original empirical studies from the overall literature set (Dataset A) | Studies involving an exercise‑related intervention in participants with rheumatoid arthritis and reporting at least one health‑related outcome; includes RCTs, quasi‑experimental, and pre‑post controlled trials | To describe the breadth of intervention research and support structured full‑text coding | Descriptive classification of intervention types (9 exercise modalities + non‑exercise co‑interventions), study designs, and outcome domains |
| Dataset C (Evidence‑matrix dataset) | 54 | Subset of original intervention studies from Dataset B with sufficiently explicit exercise prescriptions | Studies reporting a clearly classifiable single‑modality exercise protocol with extractable protocol elements (type, frequency, intensity, duration) and outcomes assignable to predefined domains; multimodal interventions with inseparable components excluded | To construct the modality‑by‑outcome evidence map and evaluate directional patterns across outcome domains | Bubble evidence matrix (Fig. 10) of exercise modalities versus clinical outcome domains |
| RCT subset | 49 | Subset of Dataset C | Randomized controlled trials with a sufficiently standardized exercise intervention protocol | To summarize trial‑design maturity and contextualize the strength of structured intervention evidence | RCT characteristic summary presented descriptively (Supplementary Table S2) |
