## Supplementary materials including additional figures and tables for "Exercise Interventions for Rheumatoid Arthritis: A Sequential Bibliometric and Content Analysis — An Evidence Mapping Study": Supplementary Table S2 .docx

Supplementary Table S2 Key design characteristics of included RCTs stratified by exercise modality

| Modality Category | Number of RCTs | Median Sample Size (IQR) | Median Intervention Duration, Weeks (IQR) | Outcome Consistency (% of studies reporting improvement in primary outcome) |
| --- | --- | --- | --- | --- |
| Behavioural/psychological | 11 | 122 (78–186) | 12 (8–24) | 72.7% |
| Aerobic training | 8 | 56 (38–89) | 12 (8–16) | 87.5% |
| Resistance training | 7 | 48 (32–72) | 12 (8–16) | 85.7% |
| Combined multimodal | 6 | 62 (44–95) | 12 (8–16) | 83.3% |
| Mind‑body (yoga/tai chi) | 5 | 52 (36–78) | 10 (8–12) | 60.0% |
| Hand/local exercises | 4 | 42 (30–58) | 4 (4–8) | 100.0%* |
| Aquatic exercise | 2 | 32 (26–38) | 12 (8–12) | 50.0% |
| HIIT | 2 | 40 (34–46) | 10 (8–12) | 100.0% |
| Whole‑body vibration | 1 | 24 | 8 | 0.0% |
| Other/novel | 3 | 30 (24–42) | 8 (6–12) | 33.3% |

*100% consistency based on 4 RCTs; total sample size across these RCTs was small (n=30–58 per study).

Abbreviations:HIIT, high‑intensity interval training; IQR, interquartile range; RCT, randomized controlled trial.
