## Supplementary materials including additional figures and tables for "Exercise Interventions for Rheumatoid Arthritis: A Sequential Bibliometric and Content Analysis — An Evidence Mapping Study": Supplementary Table S3 .docx

Supplementary Table S3 Sensitivity analysis of classification thresholds (≥3, ≥5, ≥8 studies)

| Modality | Outcome Domain | Studies (n) | Classification (≥3) | Classification (≥5) | Classification (≥8) | Stability |
| --- | --- | --- | --- | --- | --- | --- |
| Aerobic training | Disease activity | 17 | Consistent | Consistent | Consistent | Stable |
| Aerobic training | Physical function | 15 | Consistent | Consistent | Consistent | Stable |
| Aerobic training | Fatigue | 12 | Consistent | Consistent | Consistent | Stable |
| Aerobic training | Quality of life | 11 | Consistent | Consistent | Consistent | Stable |
| Aerobic training | Pain | 9 | Consistent | Consistent | Consistent | Stable |
| Aerobic training | Cardiovascular risk | 4 | Consistent | Insufficient | Insufficient | Threshold‑sensitive |
| Resistance training | Muscle strength | 14 | Consistent | Consistent | Consistent | Stable |
| Resistance training | Physical function | 11 | Consistent | Consistent | Consistent | Stable |
| Resistance training | Grip strength | 9 | Consistent | Consistent | Consistent | Stable |
| Resistance training | Disease activity | 8 | Consistent | Consistent | Consistent | Stable |
| Combined aerobic + resistance | Morning stiffness | 9 | Consistent | Consistent | Consistent | Stable |
| Combined aerobic + resistance | Functional capacity | 8 | Consistent | Consistent | Consistent | Stable |
| Mind‑body | Depression/anxiety | 8 | Inconsistent | Inconsistent | Inconsistent | Stable |
| Mind‑body | Quality of life | 7 | Inconsistent | Inconsistent | Insufficient | Threshold‑sensitive |
| Mind‑body | Fatigue | 6 | Inconsistent | Inconsistent | Insufficient | Threshold‑sensitive |
| Hand exercise | Grip strength | 6 | Consistent | Consistent | Insufficient | Threshold‑sensitive |
| HIIT | Cardiorespiratory fitness | 4 | Consistent | Insufficient | Insufficient | Threshold‑sensitive |
| Aquatic exercise | Pain | 3 | Consistent | Insufficient | Insufficient | Threshold‑sensitive |
| Whole‑body vibration | Any outcome | 1–2 | Insufficient | Insufficient | Insufficient | Stable |

Definition of stability:>90% of modality–outcome pairs remained in the same classification category (consistent/mixed/insufficient) across all three thresholds.
