## Supplementary materials including additional figures and tables for "Exercise Interventions for Rheumatoid Arthritis: A Sequential Bibliometric and Content Analysis — An Evidence Mapping Study": Supplementary Table S4.docx

Supplementary Table S4 Citation burst terms with strength values (top 15)

| Term | Strength | Begin | End | 2016–2025 |
| --- | --- | --- | --- | --- |
| patient perspective | 4.82 | 2016 | 2019 | ██████████ |
| inflammatory arthritis | 4.31 | 2017 | 2020 | ██████████ |
| oxidative stress | 3.95 | 2017 | 2019 | ████████ |
| cardiovascular risk | 3.87 | 2016 | 2019 | ██████████ |
| exercise safety | 3.62 | 2016 | 2018 | ████████ |
| fatigue | 4.56 | 2020 | 2022 | ████████ |
| cardiorespiratory fitness | 3.78 | 2021 | 2023 | ████████ |
| patient‑reported outcomes | 3.54 | 2021 | 2023 | ████████ |
| aerobic exercise | 3.41 | 2022 | 2024 | ████████ |
| systematic review | 3.22 | 2022 | 2025 | ██████████ |
| quality of life | 4.03 | 2023 | 2025 | ████████ |
| mobile applications | 3.67 | 2023 | 2025 | ████████ |
| hand function | 3.15 | 2023 | 2025 | ████████ |
| chronic pain | 2.98 | 2023 | 2025 | ████████ |
| patient involvement | 2.84 | 2024 | 2025 | ██████ |

Legend:█ Each block represents approximately 0.4 years; the length of the block bar reflects the burst duration of each term. The duration in years can be calculated by subtracting the "Begin" year from the "End" year and adding 1. Strength = citation burst intensity (CiteSpace, default γ=1.0).
