## Supplementary materials including additional figures and tables for "Exercise Interventions for Rheumatoid Arthritis: A Sequential Bibliometric and Content Analysis — An Evidence Mapping Study": Supplementary Table S5 .docx

Supplementary Table S5 Clinical outcome evidence summary (full)

| Outcome Domain | Total Studies Reporting | % Improvement | Key Consistency Pattern |
| --- | --- | --- | --- |
| Disease activity (DAS28) | 47 | 80.9% | Aerobic ≥85%; mixed modalities lower; exercise generally does not worsen disease activity |
| Pain (VAS) | 42 | 78.6% | Relatively consistent across modalities; moderate effect pattern |
| Morning stiffness | 18 | 77.8% | Combined aerobic+resistance strongest; water‑based exercise promising but limited evidence |
| Physical function (6MWT) | 28 | 82.1% | Aerobic, resistance, combined ≥80%; supports role of structured exercise in preserving mobility |
| Muscle strength | 27 | 88.9% | Resistance strongest (100%); aligns with physiological expectations |
| Grip strength/hand function | 20 | 100% | Hand exercise most consistent; highly consistent benefits even with short intervention duration (median 4 weeks) |
| Body composition | 12 | 58.3% | Inconsistent; aerobic+diet strongest; exercise alone effect unclear; limited long‑term data |
| Fatigue | 39 | 71.8% | Aerobic 85%; mind‑body 60‑70%; some studies fail to reach MCID |
| Depression/anxiety | 22 | 77.3% | Mind‑body and mHealth strongest; suggests psychophysiological effect |
| Quality of life | 35 | 77.1% | Aerobic and mixed strongest; moderate effect size (SMD ~0.50) |
| Cardiovascular risk | 15 | 66.7% | Evidence‑sparse (≤5/modality); dose‑response trend suggested but understudied |
| Vascular function | 8 | 75.0% | Positive signal but limited evidence; long‑term benefits suggested |
| Inflammatory markers | 24 | 70.8% | Mixed; some reduction in TNF‑α, IL‑6 reported; inconsistent across modalities |

Abbreviations:6MWT, 6‑minute walk test; DAS28, Disease Activity Score in 28 joints; MCID, minimal clinically important difference; SMD, standardised mean difference; TNF‑α, tumour necrosis factor‑alpha; VAS, visual analogue scale.
