## Supplementary materials including additional figures and tables for "Exercise Interventions for Rheumatoid Arthritis: A Sequential Bibliometric and Content Analysis — An Evidence Mapping Study": Supplementary Table S6.docx

Supplementary Table S6. Study characteristics of the 49 randomized controlled trials included in the content analysis, grouped by exercise modality.

| Exercise modality | Number of studies | Sample size, median (IQR) | Intervention duration (weeks), median (IQR) | Outcome consistency (%) |
| --- | --- | --- | --- | --- |
| Behavioral / psychological | 11 | 122 (72–172) | 16 (8–24) | 72.7 |
| Other* | 10 | 100 (47–152) | 12 (10–16) | 90.0 |
| Aerobic | 7 | 51 (33–77) | 14 (12–24) | 85.7 |
| Hand / local | 6 | 34 (22–45) | 4 (4–12) | 100 |
| Multimodal (diet + exercise) \| | 4 | 95 (74–117) | 16 (12–24) | 100 |
| Resistance | 4 | 75 (58–188) | 14 (12–16) | 100 |
| Mind–body | 4 | 64 (60–68) | 12 (12–12) | 100 |
| Blood flow restriction | 2 | 33 (25–41) | 12 (12–12) | 100 |
| Aquatic | 1 | 133 | 16 | 100 |

*Other includes whole‑body vibration, Argentine tango, EMG biofeedback, single‑bout acute exercise, etc.IQR: interquartile range.Outcome consistency: proportion of studies reporting improvement (“good” or “partial good”) in the primary outcome.
