## Supplementary materials including additional figures and tables for "Exercise Interventions for Rheumatoid Arthritis: A Sequential Bibliometric and Content Analysis — An Evidence Mapping Study": Table 1 .docx

Table 1 Top 10 countries by publication output and centrality on exercise interventions in rheumatoid arthritis (2016‑2025)

| Rank | Count | Centrality | Typical year | Countries |
| --- | --- | --- | --- | --- |
| 1 | 70 | 0.77 | 2016 | USA |
| 2 | 38 | 0.1 | 2016 | BRAZIL |
| 3 | 33 | 0.19 | 2016 | SWEDEN |
| 4 | 33 | 0.06 | 2016 | UK |
| 5 | 27 | 0.17 | 2019 | CHINA |
| 6 | 26 | 0.03 | 2016 | TURKEY |
| 7 | 21 | 0.17 | 2016 | NETHERLANDS |
| 8 | 18 | 0.09 | 2016 | DENMARK |
| 9 | 17 | 0.42 | 2016 | CANADA |
| 10 | 15 | 0.25 | 2016 | FRANCE |
