## Supplementary materials including additional figures and tables for "Exercise Interventions for Rheumatoid Arthritis: A Sequential Bibliometric and Content Analysis — An Evidence Mapping Study": Table 2 .docx

Table 2 Top 10 institutions by publication output on exercise interventions in rheumatoid arthritis (2016‑2025)

| Rank | Count | Centrality | Typical year | Institutions |
| --- | --- | --- | --- | --- |
| 1 | 5 | 0 | 2016 | Leiden University Medical Center |
| 2 | 5 | 0.01 | 2016 | University of Copenhagen |
| 3 | 4 | 0 | 2016 | Copenhagen Center for Arthritis Research |
| 4 | 3 | 0 | 2018 | Arai |
| 5 | 3 | 0 | 2021 | Applied Physiology and Nutrition Research Group |
| 6 | 3 | 0 | 2019 | Lange |
| 7 | 3 | 0 | 2016 | Demmelmaier |
| 8 | 3 | 0 | 2020 | Lim |
| 9 | 3 | 0 | 2017 | Fenton |
| 10 | 3 | 0 | 2016 | Rheumatology Division |
