## Supplementary materials including additional figures and tables for "Exercise Interventions for Rheumatoid Arthritis: A Sequential Bibliometric and Content Analysis — An Evidence Mapping Study": Table 3 .docx

Table 3. Top 13 authors by publication output on exercise interventions in rheumatoid arthritis (2016‑2025)

| Rank | Count | Authors | Country |
| --- | --- | --- | --- |
| 1 | 12 | Hamilton, Roschel | Brazil |
| 2 | 11 | D, Kitas George | United Kingdom |
| 3 | 11 | Bruno, Gualano | Brazil |
| 4 | 9 | S, Metsios George | United Kingdom |
| 5 | 9 | Kaisa, Mannerkorpi | Sweden |
| 6 | 8 | L, Duda Joan | United Kingdom |
| 7 | 8 | M, Fenton Sally A | United Kingdom |
| 8 | 8 | Appel, Esbensen Bente | Denmark |
| 9 | 7 | Alexander, Fraser | Ireland |
| 13 | 6 | Inger, Gjertsson | Sweden |
| 13 | 6 | Elvira, Lange | Sweden |
| 13 | 6 | Infante, Smaira Fabiana | Brazil |
| 13 | 6 | C, Rouse Peter | United Kingdom |
