## Supplementary materials including additional figures and tables for "Exercise Interventions for Rheumatoid Arthritis: A Sequential Bibliometric and Content Analysis — An Evidence Mapping Study": Table 4 .docx

Table 4. Top 10 co‑cited journals by citation frequency and their impact factors on exercise interventions in rheumatoid arthritis (2016‑2025),Rank 9 denotes a tie for 10th place.

| Rank | Cite Sources | Articles | Impact Factor(2023) |
| --- | --- | --- | --- |
| 1 | ANN RHEUM DIS | 233 | 20.3 |
| 2 | RHEUMATOLOGY | 219 | 4.7 |
| 3 | ARTHRIT CARE RES | 202 | 4.7 |
| 4 | Arthritis & Rheumatology | 169 | 10.9 |
| 5 | J RHEUMATOL | 161 | 3.9 |
| 6 | CLIN RHEUMATOL | 148 | 2.9 |
| 7 | RHEUMATOL INT | 143 | 3.2 |
| 8 | ARTHRITIS RES THER | 140 | 4.9 |
| 9 | MED SCI SPORT EXER | 121 | 3.9 |
| 9 | LANCET | 106 | 88.5 |
