## Supplementary materials including additional figures and tables for "Exercise Interventions for Rheumatoid Arthritis: A Sequential Bibliometric and Content Analysis — An Evidence Mapping Study": Table 5.docx

Table 5. Evidence volume and directional consistency of exercise modalities for rheumatoid arthritis.

| Exercise Modality | Evidence Volume | Directional Consistency | Main Supported Outcomes | Identified Research Gaps |
| --- | --- | --- | --- | --- |
| Aerobic training | High (≥15 studies) | High (≥85%), | DAS28, VO₂peak, fatigue, quality of life | Long‑term adherence (>1 yr); optimal dosing |
| Resistance training | High (≥15 studies) | High (≥85%) | Muscle strength, lean mass, 6MWT, grip strength | Dosing parameters; long‑term follow‑up |
| Combined aerobic + resistance | Moderate (≥10 studies) | Moderate (≥80%) | Morning stiffness, functional capacity | Protocol standardisation |
| Mind–body (yoga, tai chi) | Moderate (16 studies) | Low (60‑70%) | Depression, anxiety, quality of life | Standardised protocols; larger confirmatory trials |
| Water-based exercise | Low (≤5 studies) | Low | Pain, morning stiffness | Larger RCTs |
| HIIT / BFRT | Very low (≤5 studies) | Very low | Fatigue (HIIT), muscle strength (BFRT) | Safety/feasibility; dose‑finding |
| Hand / local exercises | Low (≤5 per outcome pair) | High (100% in RCTs)* | Grip strength, dexterity | Larger confirmatory trials; home‑based delivery |
| Whole‑body vibration | Very low (≤2) | Insufficient | — | Pilot sham‑controlled RCTs |
