## Supplementary figures and images for "Exercise Interventions for Rheumatoid Arthritis: A Sequential Bibliometric and Content Analysis — An Evidence Mapping Study"

### Fig S1.png

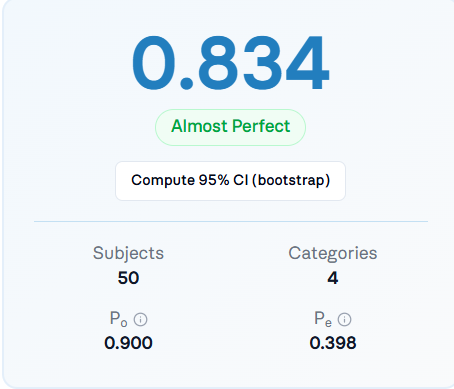

### Fig. 1.jpg

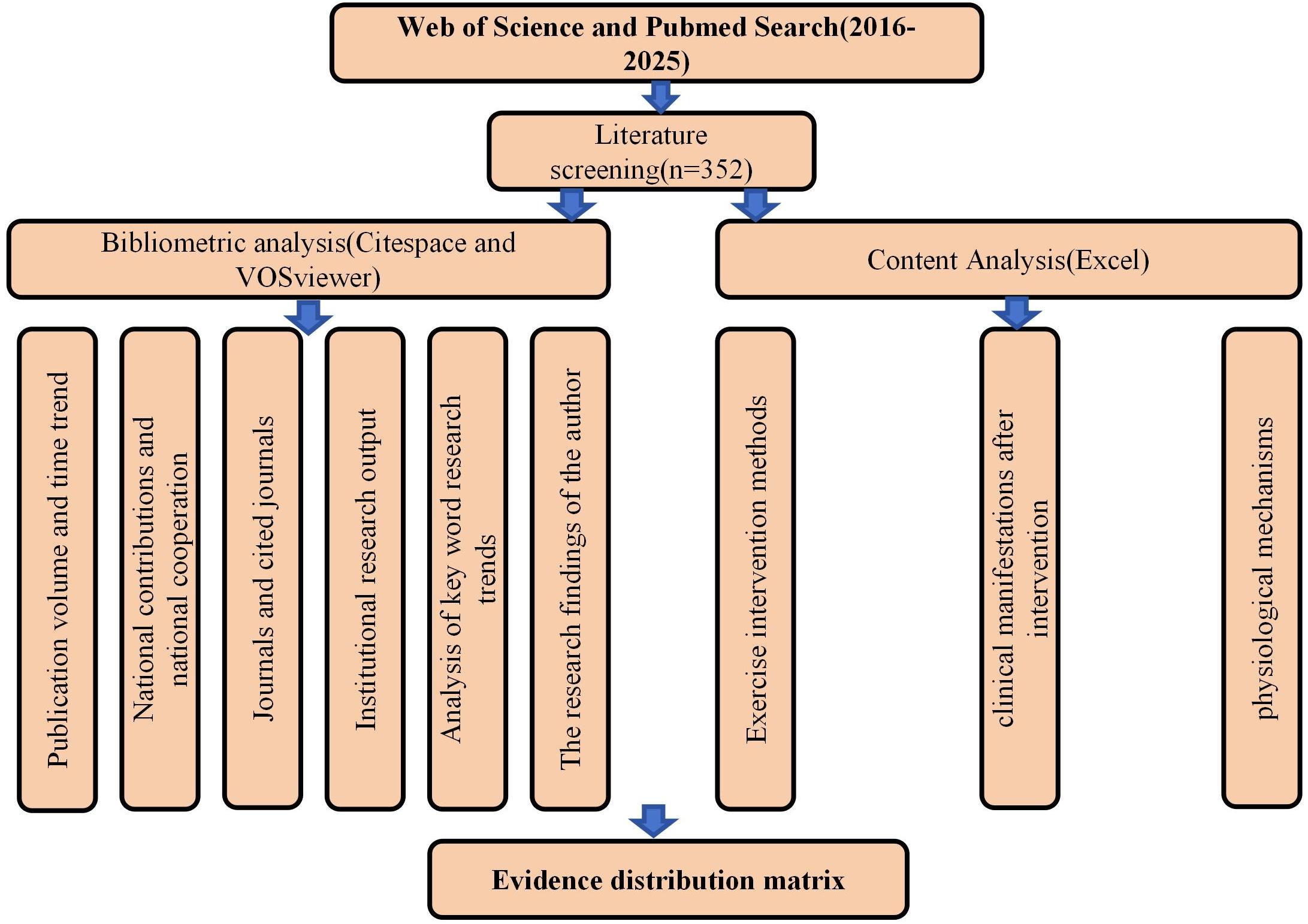

### Fig. 2.jpg

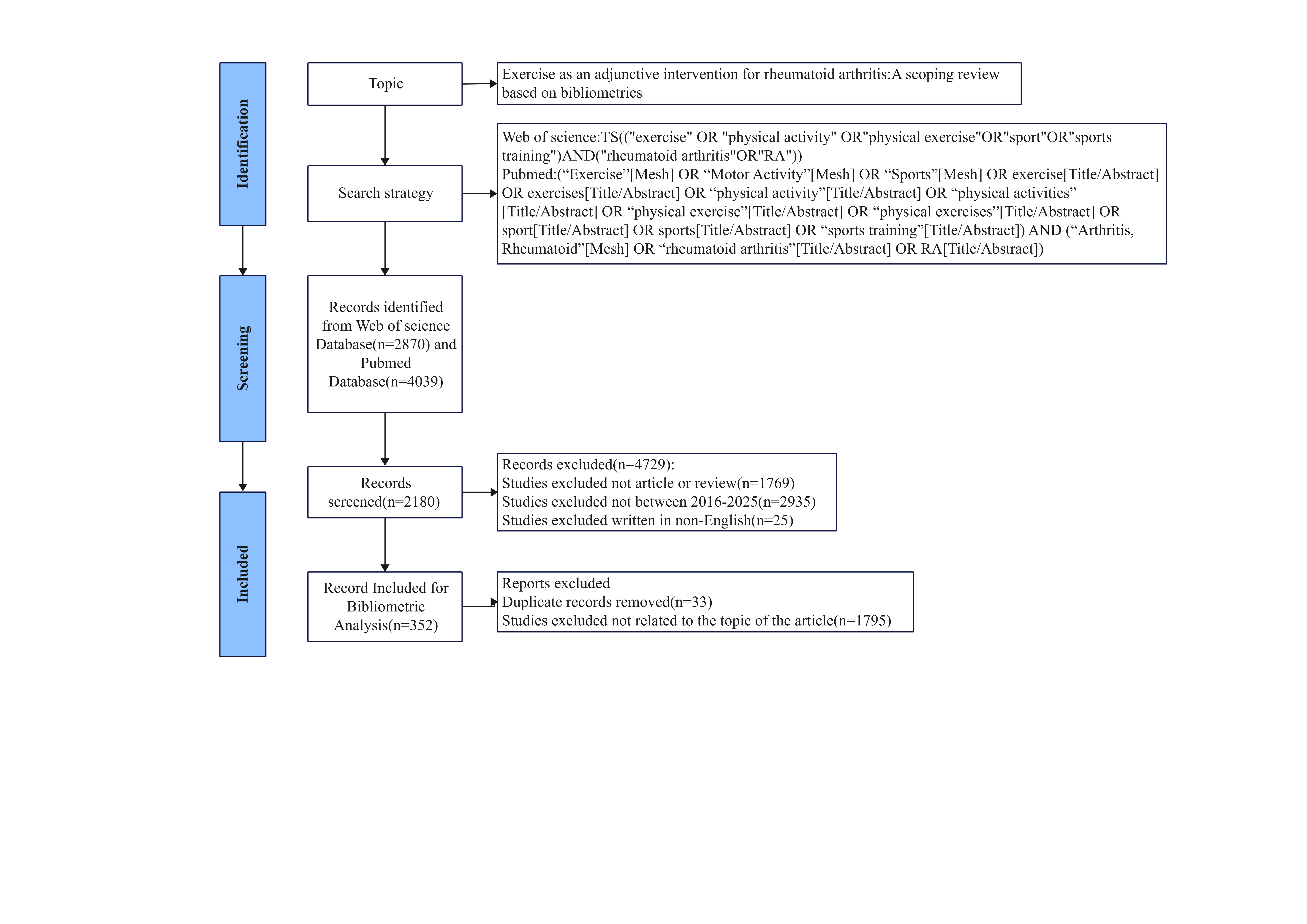

### Fig. 3.png

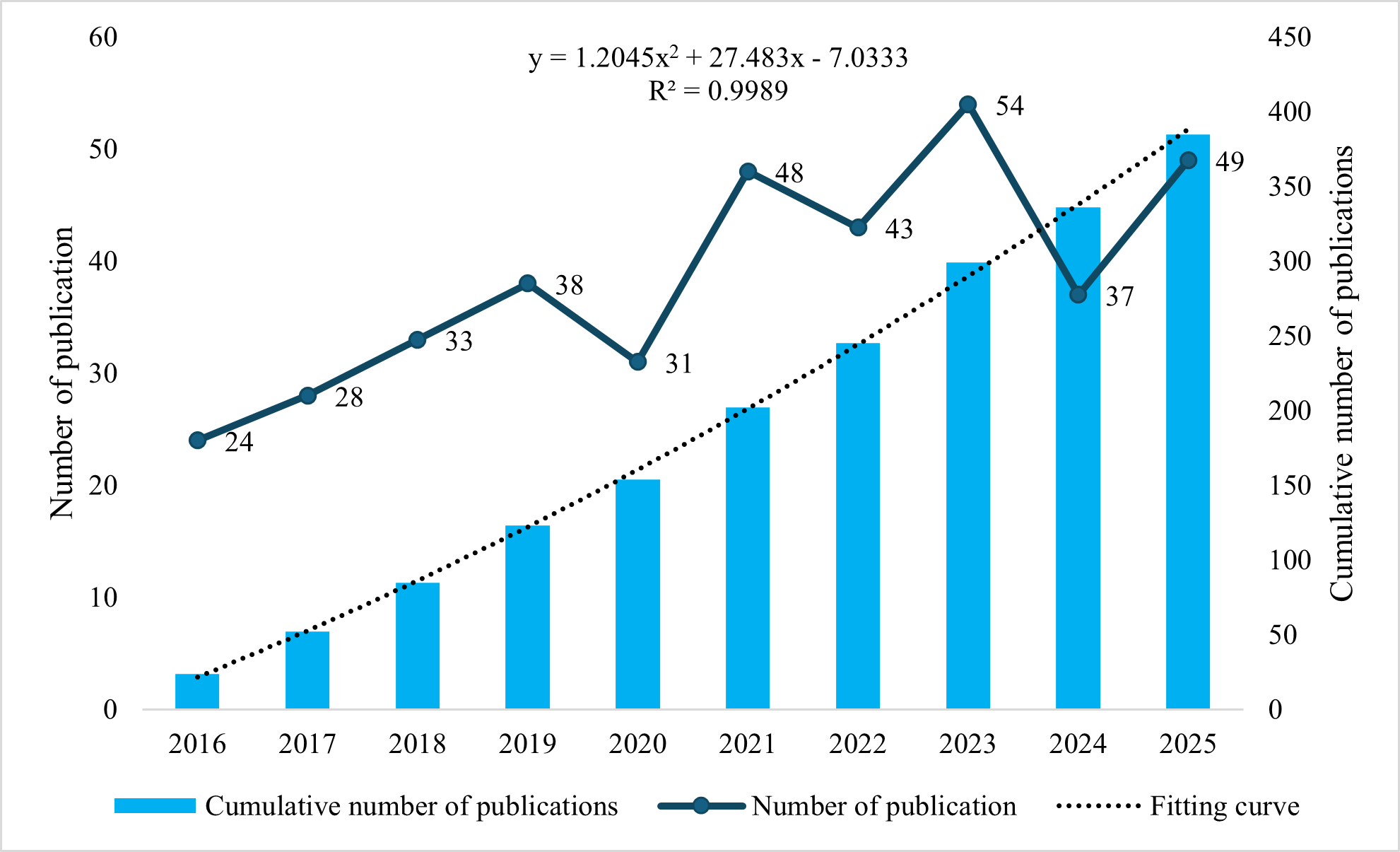

### Fig. 4.png

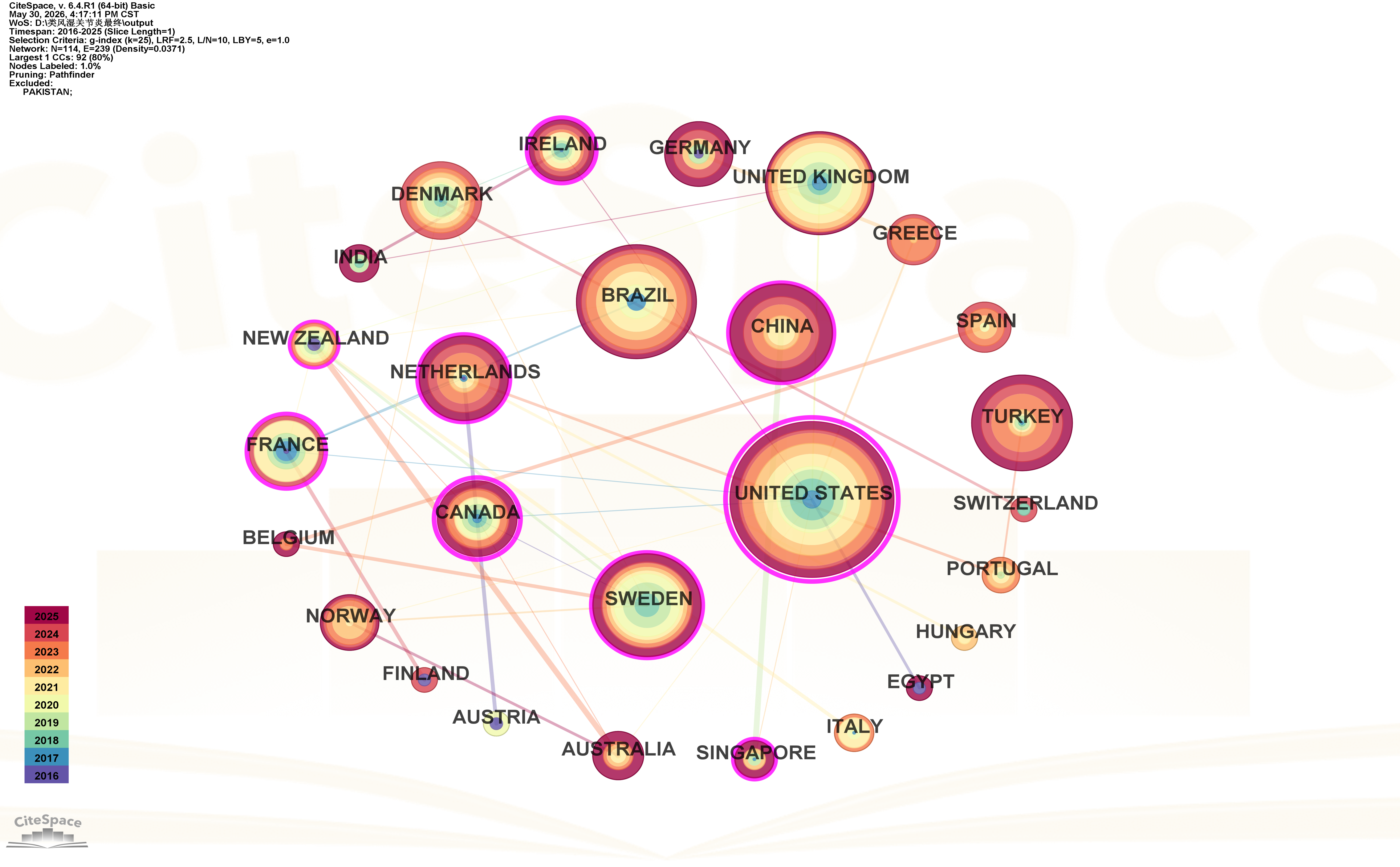

### Fig. 5.png

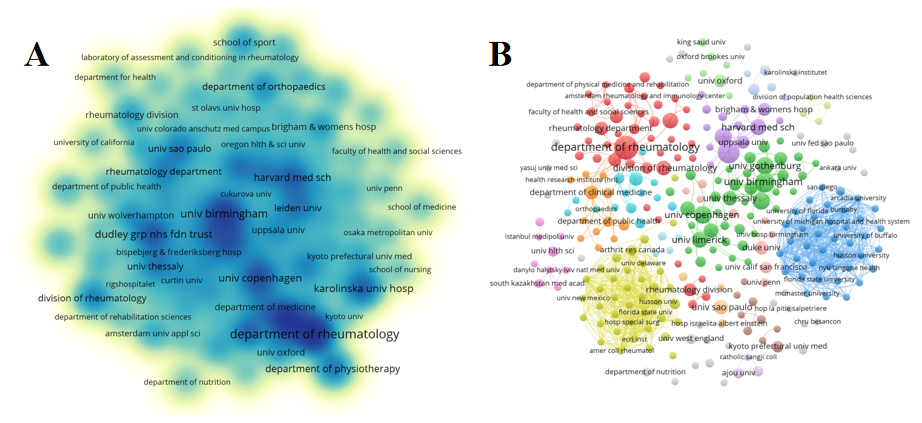

### Fig. 6.png

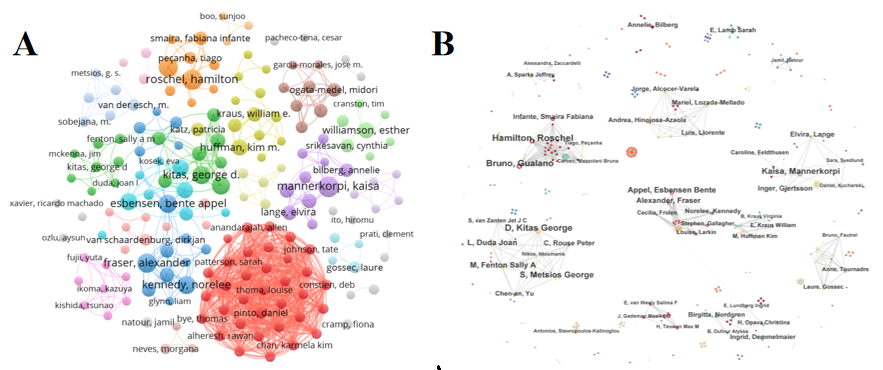

### Fig. 7.png

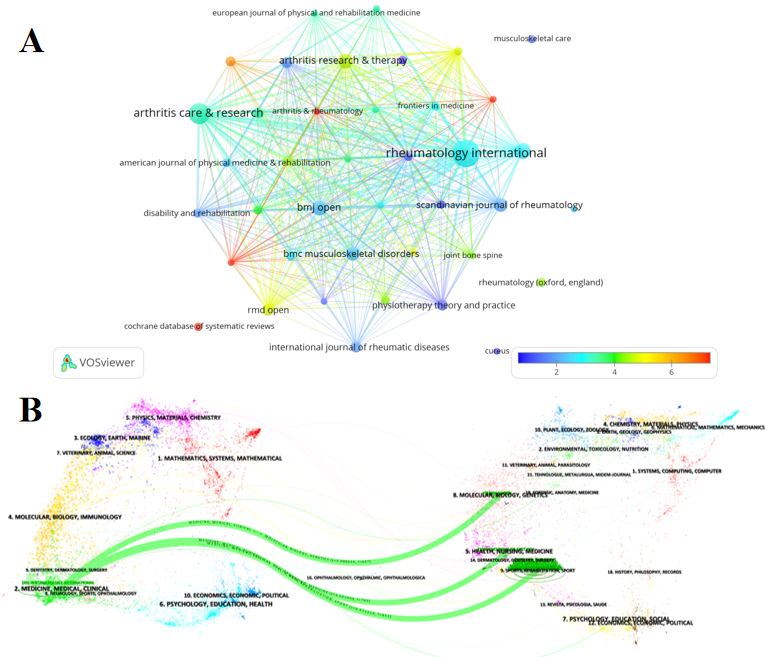

### Fig. 8.png

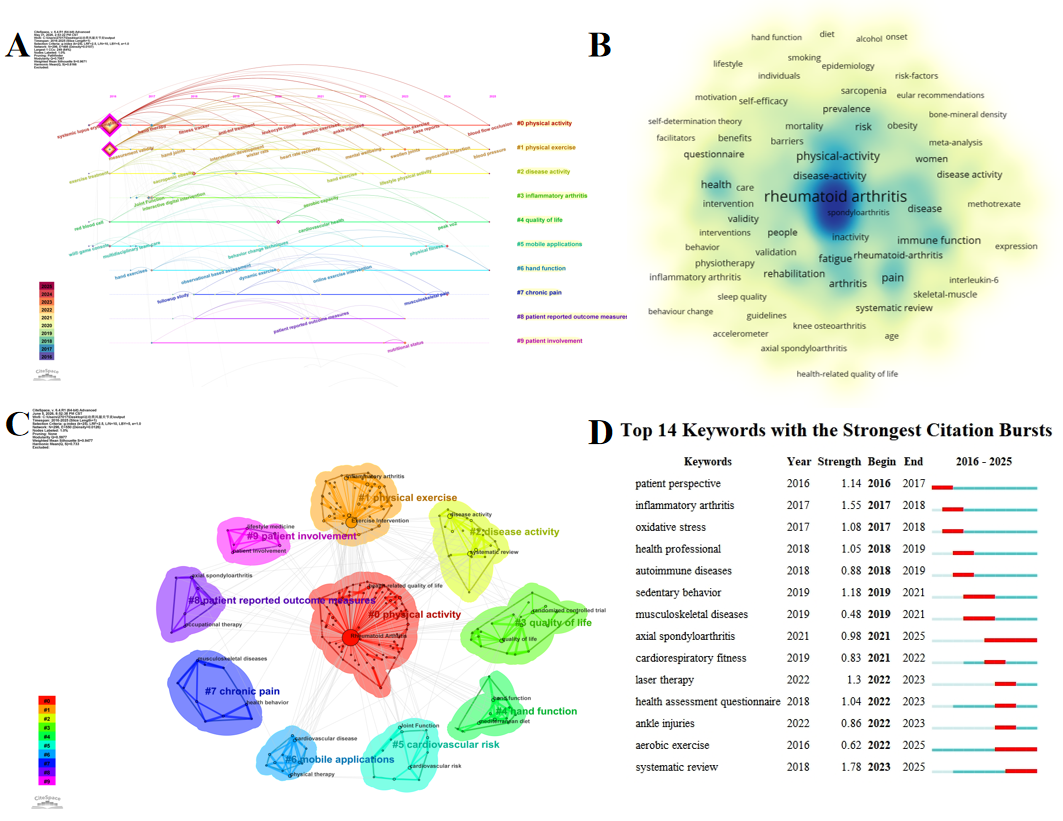

### Fig. 9.png

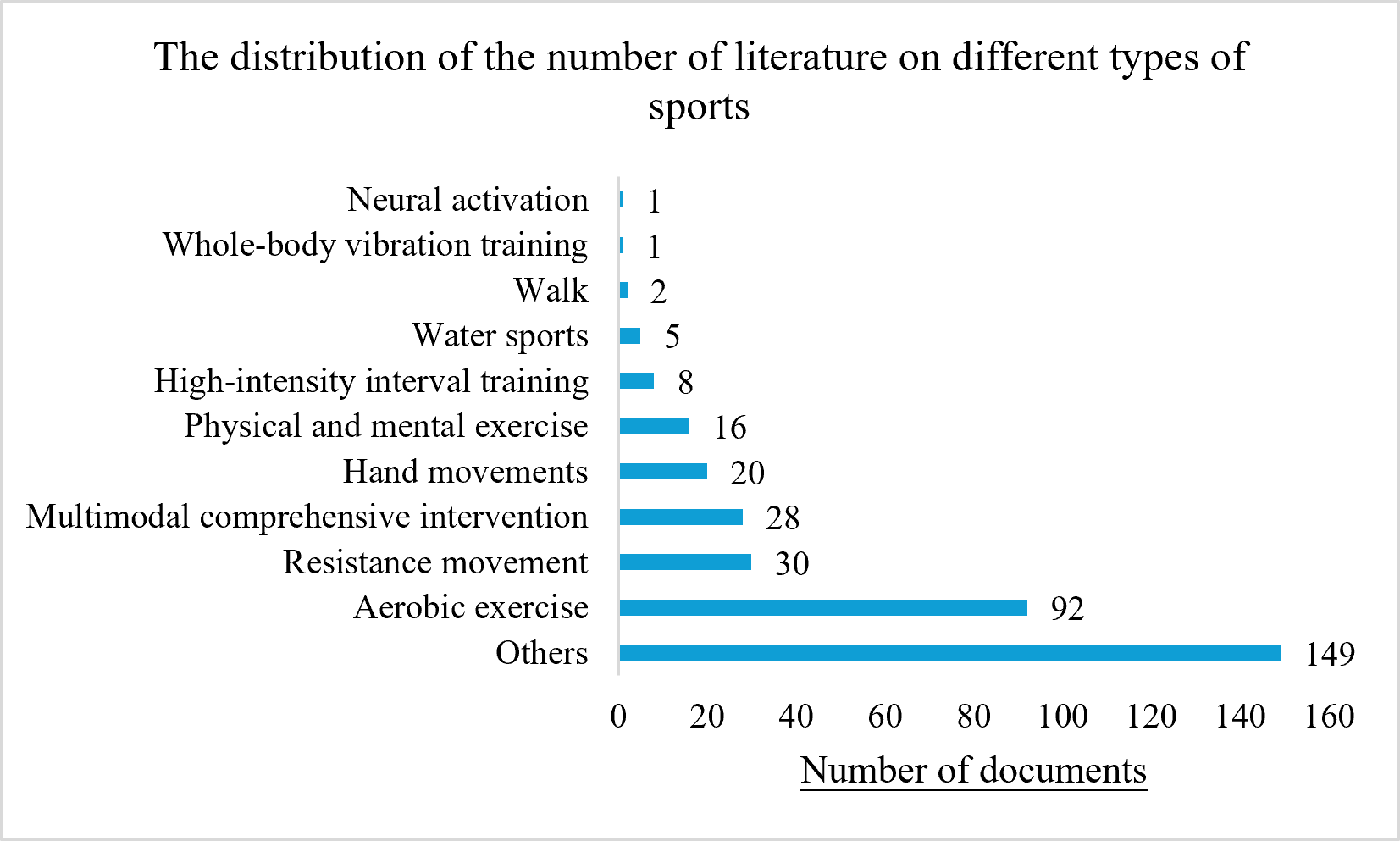

### Fig. 10.png

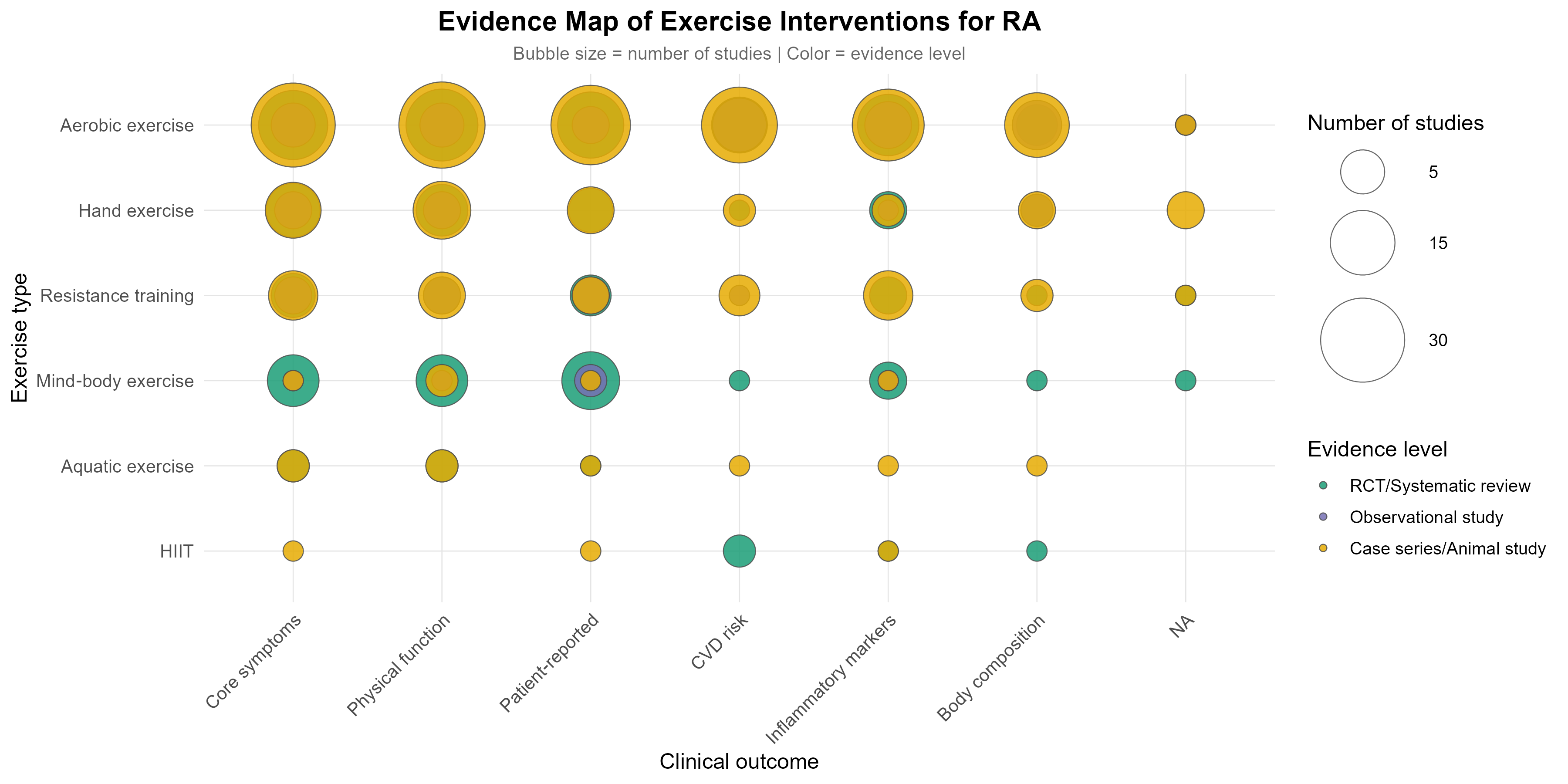
